# Stratification of Alzheimer’s Disease Patients Using Knowledge-Guided Unsupervised Latent Factor Clustering with Electronic Health Record Data

**DOI:** 10.1101/2024.12.23.24319588

**Authors:** Linshanshan Wang, Shruthi Venkatesh, Michele Morris, Mengyan Li, Ratnam Srivastava, Shyam Visweswaran, Oscar Lopez, Zongqi Xia, Tianxi Cai

## Abstract

Prognostication for people with Alzheimer’s disease (AD) at the point of care could improve clinical management. Applying a novel unsupervised latent factor clustering approach guided by knowledge graph embeddings of relevant clinical features from electronic health records, we stratified 16,411 AD patients into two groups at diagnosis and prognosticated their risk of AD-related outcomes (*i.e.,* nursing home admission, mortality), adjusting for baseline confounders. To reflect real-world evolution in clinical trajectories, we updated patient stratification for 12,606 AD patients remaining at risk 1-year post-diagnosis and repeated prognostication. At both timepoints, one group had a higher nursing home admission risk and exhibited characteristics suggesting greater symptom burden, but the mortality risk remained comparable between groups. This study supports that patient stratification can enable outcome prognosis for AD patients. While baseline prognostication can guide early treatment and tailored management, dynamic prognostication may inform more timely interventions to improve long-term outcomes.

## INTRODUCTION

Alzheimer’s disease (AD) is the leading cause of dementia and neurological disability in the aging population, causing high societal burdens.^1–3^ As promising treatment options emerge and given their potential tradeoff (*e.g.,* adverse events, cost),^4,5^ there is a heightened interest in patient stratification and prognostication at the point of care. Multifactorial contributors of the varying patient trajectories pose a major challenge to effective clinical prediction.^6–11^

The increasing availability of large-scale clinical datasets and advances in machine learning (ML) algorithms enable data-driven investigations of heterogeneity in AD clinical trajectories. Unsupervised ML techniques such as clustering can stratify AD patient populations into more homogeneous groups based on latent clinical features relevant to AD. Iterative patient stratifications using baseline and subsequently evolving clinical features can inform contributors of differential risks in clinical trajectories, and pragmatically improve clinical prediction and ultimately treatment guidance.^12^

Prior AD clustering studies used genetics,^13^ biomarkers,^14–16^ neuropathology neuroimaging,^17–22^ and neuropsychological data.^16,23,24^ However, these studies were limited in follow-up duration, scalability, generalizability and practicality. Electronic health records (EHRs) contain large-scale, real-world, longitudinal data for clinical investigations of AD. Several studies have leveraged EHR data to cluster AD patients for prediction.^25–29^ Two studies clustered patients based on symptomatic presentation and comorbid conditions preceding AD onset for predicting time to AD onset.^25,26^ Two other studies clustered patients based on pre-selected lists of features for predicting AD outcomes such as mortality,^27,28^ but pre-selected clinical features lack scalability and generalizability. Patient clustering can be enhanced by automated selection of clinically informative features based on data-derived prior knowledge that incorporates complementary features embedded in the EHR such as comorbid conditions, medication prescriptions, test results, and clinical narratives.

Here, we aim to stratify AD patients at and one year after diagnosis to prognosticate readily ascertainable AD-related outcomes of nursing home admission and mortality, using baseline clinical features. Leveraging previously trained knowledge graph embeddings of EHR concepts, we automated the selection of AD-relevant clinical features to guide a latent class model of patient clustering.^30^ Patient cluster memberships based on clinical profile before AD diagnosis and updated cluster membership one year post-diagnosis predicted future AD outcomes.

## METHODS

### Ethics approval

The Institutional Review Board of the University of Pittsburgh (STUDY21020153) approved the study protocol.

### AD data mart

We obtained inpatient and outpatient EHR data for years 2011 to 2022 from the UPMC healthcare system, which comprises academic and community practices in a large catchment and is representative of its broader geographic region. Codified EHR data contain demographics (*e.g.*, age, gender, race, ethnicity), diagnosis and procedure codes (*e.g.*, International Classification of Disease [ICD] code, Current Procedural Terminology [CPT] code), healthcare utilization metrics (*e.g.*, total number of ICD codes and encounter notes), medication prescriptions (*e.g.*, RxNorm codes) and laboratory test results (*e.g.*, Logical Observation Identifiers Names and Codes [LOINC] codes).^31,32^ To systematically organize diagnoses, we mapped all ICD-9 and ICD-10 codes to PheCodes,^33,34^ a standardized method that groups related ICD codes under the same diagnosis. We consolidated procedure codes (CPT) using the Clinical Classifications Software (CCS) for Services and Procedures (CCS-Services and Procedures), a tool developed by the Agency for Healthcare Research and Quality (AHRQ) that categorizes medical procedures into clinically meaningful groups.

Narrative EHR data comprise free-form text information from clinical encounters (*e.g.,* physician office visit notes, discharge summaries) that were converted to structured data by natural language processing (NLP). Using codified and narrative EHR data, we built an AD data mart comprising patients with at least one ICD code for AD or related dementia (*e.g.,* ICD-9=290.x, 294.2x, 331.0; ICD-10=F03.9x, G30.x**, S-Table 1**). To obtain standardized clinical concepts from narrative EHR data, we deployed a previously validated NLP pipeline (Narrative Information Linear Extraction [NILE]) to generate concept unique identifiers (CUIs) according to unified medical language system (UMLS).^35,36^

### AD cohort

We applied a phenotyping algorithm, Knowledge driven Online Multimodal Automated Phenotyping (KOMAP),^30^ to assign AD diagnosis status (probable or possible AD vs not AD) for all patients in the AD data mart (**Figure 1A**). We combined probable and possible AD into a single category for prediction of AD diagnosis status as they represent heterogenous presentations of AD. KOMAP is a two-step unsupervised algorithm that enables retrieval of diagnosis labels from EHR data. First, KOMAP leverages an online narrative and codified feature search engine (ONCE) powered by multi-source knowledge graph representation learning to generate a list of informative codified and narrative features relevant to AD. Second, we trained the KOMAP algorithm based on ONCE-selected features. We obtained the AD cohort by including patients assigned by KOMAP as having probable or possible AD at 90% specificity. We included patients with at least one AD PheCode and 24 months of EHR data preceding the index date (date of the first AD PheCode, which was operationally defined as the date of AD diagnosis). We excluded patients who had at least one code for nursing home admission preceding AD diagnosis. We imputed missing demographics (*e.g.,* race and ethnicity) by performing multiple imputation based on age and gender.^37,38^

**Figure 1.**
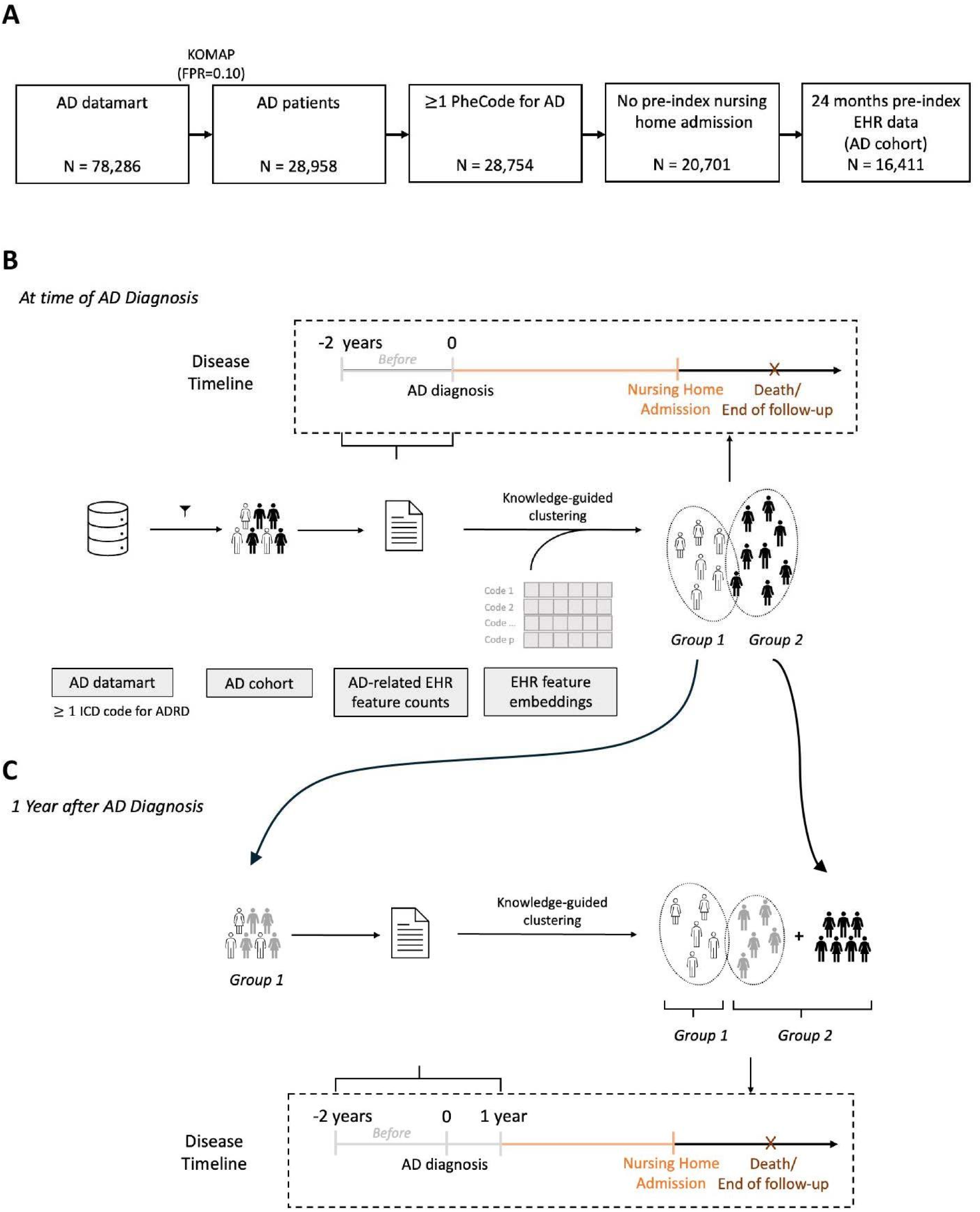
Schematic overview. (A) Inclusion criteria. (B) Study workflow.

### Clustering patients at AD diagnosis

#### Feature selection and knowledge graph embeddings

To cluster patients at AD diagnosis, we selected all AD-related codified and narrative features using ONCE and aggregated counts of EHR features for the 24 months preceding the index date.^30^ We obtained embeddings of selected concepts, denoted by *E*, through multi-source knowledge graph representation learning. This knowledge graph leverages the pre-trained ONCE embeddings of large scale EHR concepts learned from summary level EHR data at two independent healthcare systems (1,25,00,000 Veterans Health Administration patients and 60,000 Mass General Brigham Biobank patients) and the textual descriptions of the concepts embedded using CODER pre-trained language model.^39^ Importantly, this multi-source representation of EHR concepts contains rich information on the concept-concept relationships, which are generalizable across diseases and health systems.

### Identifying clusters and model specification

To identify the underlying patient clusters, we adopted the multivariate Poisson-LogNormal Mixture Model (PLMM) to jointly model the EHR feature counts. We set the number of clusters to two before fitting the model to represent a low-risk and a high-risk group, dividing patients into two groups (Group 1 and Group 2) at AD diagnosis (**S-Table 2**). We specified the model as:

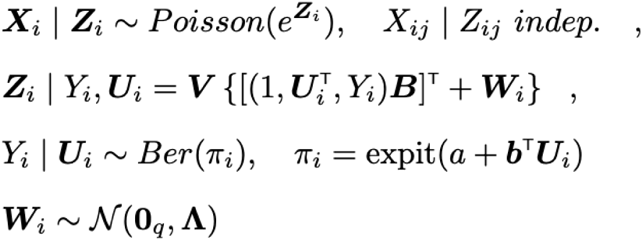

In this model, the observed variable represents the counts of *j* th feature (*e.g.,* PheCode, NLP mentions) for the *i* th patient. denotes the healthcare utilization (calculated as the total number of clinical encounters). is a latent variable denoting the two underlying patient clusters, where each cluster is assumed to have differential distributions of feature counts and downstream AD-related outcomes. represents the latent patient-level embedding for *i* th patient, with each cluster having a different mean. A graphical representation of the model is shown in **S-Figure 1A**.

Unlike previous ecological and genetics studies that use PLMM, we introduce an additional assumption: the observed EHR counts are influenced by a small number of latent factors. These latent factors capture underlying patterns of similarity (hierarchical relationships, *e.g.,* “PheCode:290”, “PheCode:290.1” and “PheCode:290.11”) and relatedness (*e.g.,* “associated with”, “may treat”, “co-occur with” etc.) among EHR features, reflecting shared relationships or common processes in the data (**S-Figure 1B**). Further, we incorporate knowledge graph embeddings, which provide rich contextual information about the relationships between concepts, to guide the identification of these factors. The latent factors serve as a bridge, summarizing complex, high-dimensional data into a simpler structure that still retains key information. Model parameters can be interpreted as the effect of the *j* th factor on cluster membership, after adjustment for healthcare utilization. The free model parameters can then be viewed as a fine-tuning parameter to allow for re-weighting of each factor based on the observed data. See supplementary methods for details on model training.

### Clustering patients one year after AD diagnosis

To illustrate the capacity of the approach to account for potential changes in patient profile that could influence trajectory, we updated the clustering model one year post-diagnosis for the patients remaining at risk (**Figure 1C**). The patients that remained at risk included those who were: (1) not admitted to nursing home, (2) not censored, (3) alive one year post-diagnosis, and (4) assigned to Group 2 at baseline. We reclustered the patients into Groups 1 and 2 leveraging 24 months of pre-index data and 12 months of post-index data. The updated Group 1 comprised patients that were reassigned from Group 2 to Group 1 one year post-diagnosis and patients in Group 1 at AD diagnosis. The updated Group 2 comprised patients that continued to be assigned to Group 2 one year post-diagnosis.

### AD-related outcomes

We assessed two AD-related outcomes as endpoints readily ascertainable from EHR data: time-to-nursing home admission and time-to-death. Admission to a nursing home, which indicates loss of functional independence, is routinely documented in clinical practice and EHR.^6,7^ We defined nursing home admission as having at least one ICD code for admission to any residential institution (*e.g.,* assisted living facility, long term care facility, end-of-life care, see **S-Table 3** for list of nursing home codes). Death status in the UPMC EHR data is linked to the social security death index. For patients that died during the follow-up period, we calculated the time-to-death from the index date. For patients that did not progress to either endpoint, we utilized the data available until the last record of clinical visit.

### Validation analyses

#### Characterizing the AD cohort

We report baseline demographics, clinical characteristics, pre-existing comorbidity burden, and prescription of AD-related medications (*e.g.,* donepezil, memantine, see **S**-Table 4 for full list of medications) during the follow-up period stratified by model-predicted clusters. We assessed the pre-existing comorbidity burden using the Elixhauser Comorbidity Index (ECI). Using the comorbidity package, we mapped the diagnosis codes for each patient to 29 pre-existing health conditions comprising the ECI.^40^ Each comorbidity in the ECI was used as an individual feature in downstream analyses. We used two-tailed t-tests to evaluate differences in baseline characteristics between clusters. To examine the differences in pre-existing comorbidity burden by predicted clusters, we calculated the proportion of patients with each comorbidity and used two-proportion Z-tests to evaluate the group difference.

### Prognostication of AD-related outcomes at time of diagnosis and 1-year post diagnosis

We examined the association between cluster membership at diagnosis and AD-related outcomes in competing risk Cox proportional hazard models. We assessed the time-to-nursing home admission alive using a competing risk model to control for death. Further, we compared the 2-year, 5-year, and 10-year rate of nursing home admission and death between clusters at diagnosis. We performed the same analyses after updating cluster membership one year post-diagnosis.

### Feature importance

We examined the salient EHR features that distinguish patients in each baseline and updated cluster. We first obtained the counts of each AD-related feature (**S-Table 5, S-**Table 6). Next, for each feature, we regressed the log-transformed (x -> log(x+1)) feature count against log-transformed healthcare utilization, and calculated the *adjusted intensity* as the mean of the residuals in each cluster. We obtained a feature importance score for each feature as the difference in adjusted intensity between the two clusters. This score reflects the difference in count frequencies between the clusters while accounting for the variation in healthcare utilization across patients.

### Software availability

The code to train and apply the PLMM is implemented in R and C++ and is publicly available at https://github.com/wanglss-2024/ADstratification, and the multi-source knowledge graph embeddings is at https://shiny.parse-health.org/ONCE/.^30^

## RESULTS

### Prediction of outcomes at AD diagnosis Demographic characteristics

The overall AD cohort comprised 16,411 patients with mean age 81.2±9.0 years at diagnosis (**Table 1**). Race and ethnicity data were missing for a small proportion of the cohort (793 / 16,411 patients). As cohort characteristics were similar with and without imputation (**S-Table 7**), we performed downstream analyses using the full AD cohort with imputed race and ethnicity.

Using the baseline clinical profile, the model clustered the AD cohort into two groups at diagnosis (Group 1: 6,734, 41.0%; Group 2: 9,677, 59.0%). Group 1 was marginally older at diagnosis (age at AD diagnosis Mean [SD]: Group 1: 81.4 [9.3], Group 2: 81.0 [8.7], *p*=.007). While the proportion of men and women was comparable between groups (Group 1: 4,314, 64.1%; Group 2: 6,245, 64.5%; *p*=.546), the proportion of non-Hispanic White individuals was higher in Group 2 (Group 1: 6,138, 91.1%; Group 2: 8,967, 92.7%; *p*=.014). The overall comorbidity burden was higher in Group 1 (ECI, Mean [SD]: Group 1: 11.3 [10.3]; Group 2: 7.5 [8.6]; *p*<.0001), with hypertension, cardiac arrhythmias, depression, and fluid and electrolyte disorders being the most prevalent comorbidities (**Table 1**, **Figure 2**).

**Figure 2.**
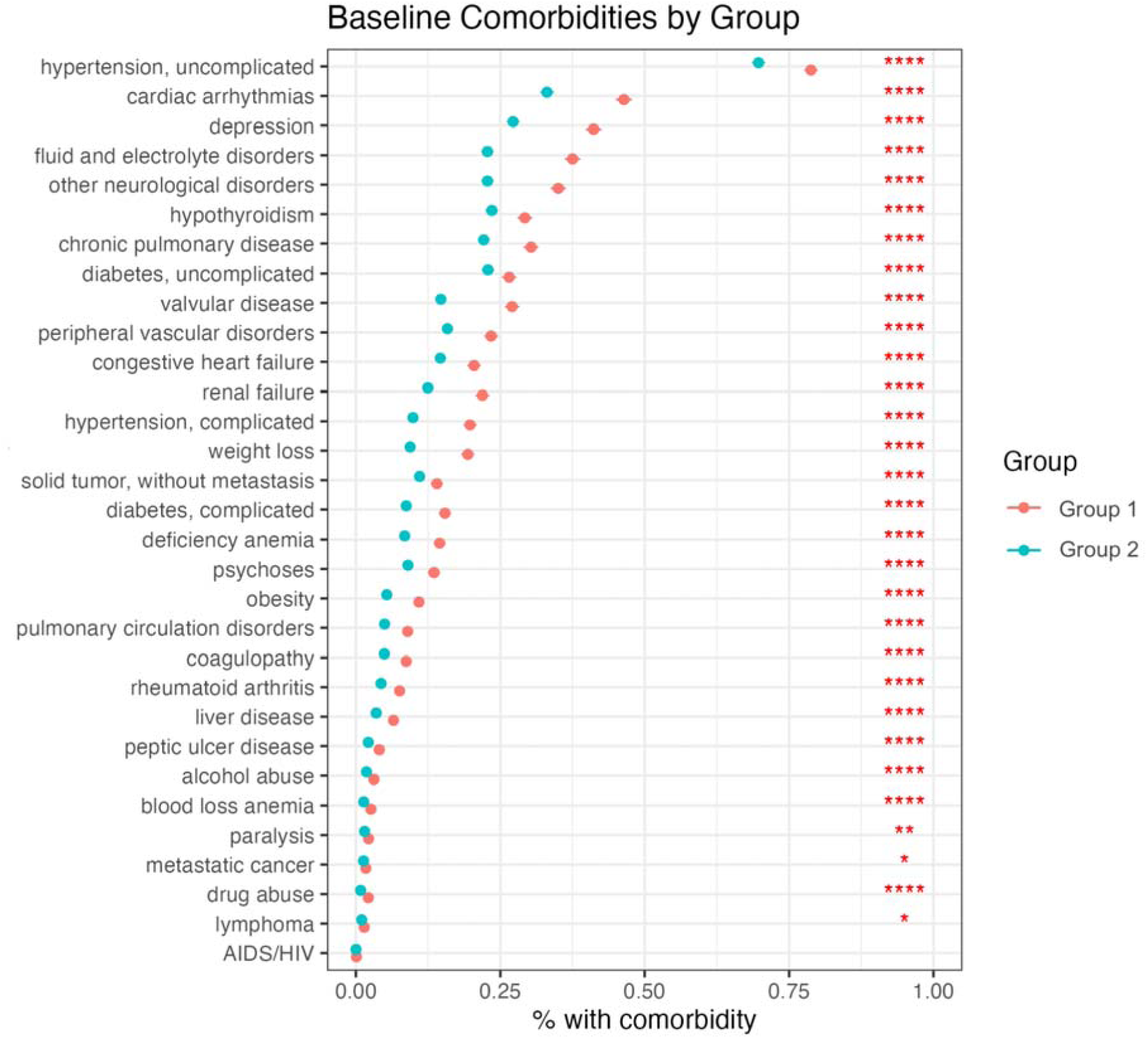
Baseline comorbidity burden of AD patient groups clustered at AD diagnosis.

In the overall AD cohort, 12,702 patients (77.4%) were prescribed AD-related medications, including 43.7% preceding diagnosis and 33.7% after diagnosis (**Table 1**). A greater proportion of patients in Group 1 were prescribed AD-related medications than Group 2 (Group 1: 47.7%; Group 2: 40.9%; *p*<.0001), suggesting higher symptom severity in Group 1. Group 1 also had a shorter follow-up duration (median [IQR]: Group 1: 62 [35, 85], Group 2: 86 [59, 125], *p*<.0001), suggesting that individuals in Group 1 progressed to the endpoints (nursing home admission or death) faster than individuals in Group 2.

**Table 1.** Demographic and clinical profile of the overall study population and patient groups clustered at AD diagnosis.

|  | Overall | Group 1 | Group 2 | P-value |
| --- | --- | --- | --- | --- |
| **Number of Patients, N (%)** | 16411 (100%) | 6734 (41.0%) | 9677 (59.0%) |  |
| **Age at AD diagnosis, mean (SD), years** | 81.2 (9.0) | 81.4 (9.3) | 81.0 (8.7) | **.007** |
| **Gender, N (%)** |  |  |  | **.546** |
| Women | 10559 (64.3%) | 4314 (64.1%) | 6245 (64.5%) |  |
| Men | 5852 (35.7%) | 2420 (35.9%) | 3432 (35.5%) |  |
| **Race/ethnicity<sup>a</sup>, N (%)** |  |  |  | **.014** |
| Non-Hispanic white | 15105 (92.0%) | 6138 (91.1%) | 8967 (92.7%) |  |
| American Indian | 8 (0.0%) | 3 (0.0%) | 5 (0.1%) |  |
| Asian | 65 (0.4%) | 32 (0.5%) | 33 (0.3%) |  |
| Black | 1183 (7.2%) | 535 (7.9%) | 648 (6.7%) |  |
| Hawaiian or Pacific Islander | 3 (0.0%) | 2 (0.0%) | 1 (0.0%) |  |
| Hispanic or Latino | 47 (0.3%) | 24 (0.4%) | 23 (0.2%) |  |
| **Elixhauser Comorbidity Index<sup>b</sup>, mean (SD)** | 9.0 (9.5) | 11.3 (10.3) | 7.5 (8.6) | **<.0001** |
| **Baseline comorbidities (top 2 prevalent), N (%)** |  |  |  | **<.0001** |
| Hypertension, uncomplicated | 12052 (73.4%) | 5305 (78.8%) | 6747 (69.7%) |  |
| Cardiac arrhythmias | 6326 (38.5%) | 3124 (46.4%) | 3202 (33.1%) |  |
| **AD-related medication<sup>c</sup>, N (%)** |  |  |  |  |
| Not prescribed | 3709 (22.6%) | 1794 (26.6%) | 3742 (38.7%) | **<.0001** |
| Prescribed | 12702 (77.4%) | 4940 (73.4%) | 5935 (61.3%) |  |
| Before AD diagnosis | 7166 (43.7%) | 3212 (47.7%) | 3954 (40.9%) | **<.0001** |
| On or after AD diagnosis | 5536 (33.7%) | 1794 (26.6%) | 3742 (38.7%) |  |
| **Follow-up months, median months (1st and 3rd quantiles)** | 78 (48, 109) | 62 (35, 85) | 86 (59, 125) | **<.0001** |

Note:

### Time to AD-related outcomes

We compared the rates of nursing home admission and mortality at the 2-year, 5-year, and 10- year milestone. While Group 1 had a higher 2-year and 5-year nursing home admission rate, the groups did not differ in the 10-year nursing home admission rate (**Table 2**). Group 1 also had a higher 2-year and 10-year mortality rate than Group 2, but there was no difference in the 5-year mortality rate between the groups.

**Table 2.** Comparison of 2-, 5- and 10-year rate of nursing home admission and death for AD patient groups clustered at AD diagnosis (N = 16,411).

|  | Nursing Home Admission |  |  | Death |  |  |
| --- | --- | --- | --- | --- | --- | --- |
|  | Group 1 | Group 2 | P-value | Group 1 | Group 2 | P-value |
| **2 years** | 31.0% (29.9%, 32.1%) | 23.5% (22.6%, 24.3%) | **<.0001** | 23.2% (22.2%, 24.3%) | 19.1% (18.3%, 19.9%) | **<.0001** |
| **5 years** | 45.5% (44.3%, 46.7%) | 40.2% (39.3%, 41.2%) | **<.0001** | 44.1% (42.9%, 45.3%) | 43.6% (42.6%, 44.6%) | .566 |
| **10 years** | 48.8% (47.6%, 50.0%) | 47.5% (46.5%, 48.5%) | .0919 | 49.6% (48.4%, 50.8%) | 61.6% (60.6%, 62.6%) | **<.0001** |

In survival analysis after adjusting for baseline covariates (age, gender, race, ethnicity, healthcare utilization, and comorbidities), Group 2 had a significantly lower risk of nursing home admission (HR [95% CI]=0.804 [0.765, 0.844], *p*<.001, **Figure 3**) than Group 1. However, the risk of mortality was comparable among the groups (HR [95% CI]=1.008 [0.963, 1.056], *p*=0.733) after covariate adjustment.

**Figure 3.**
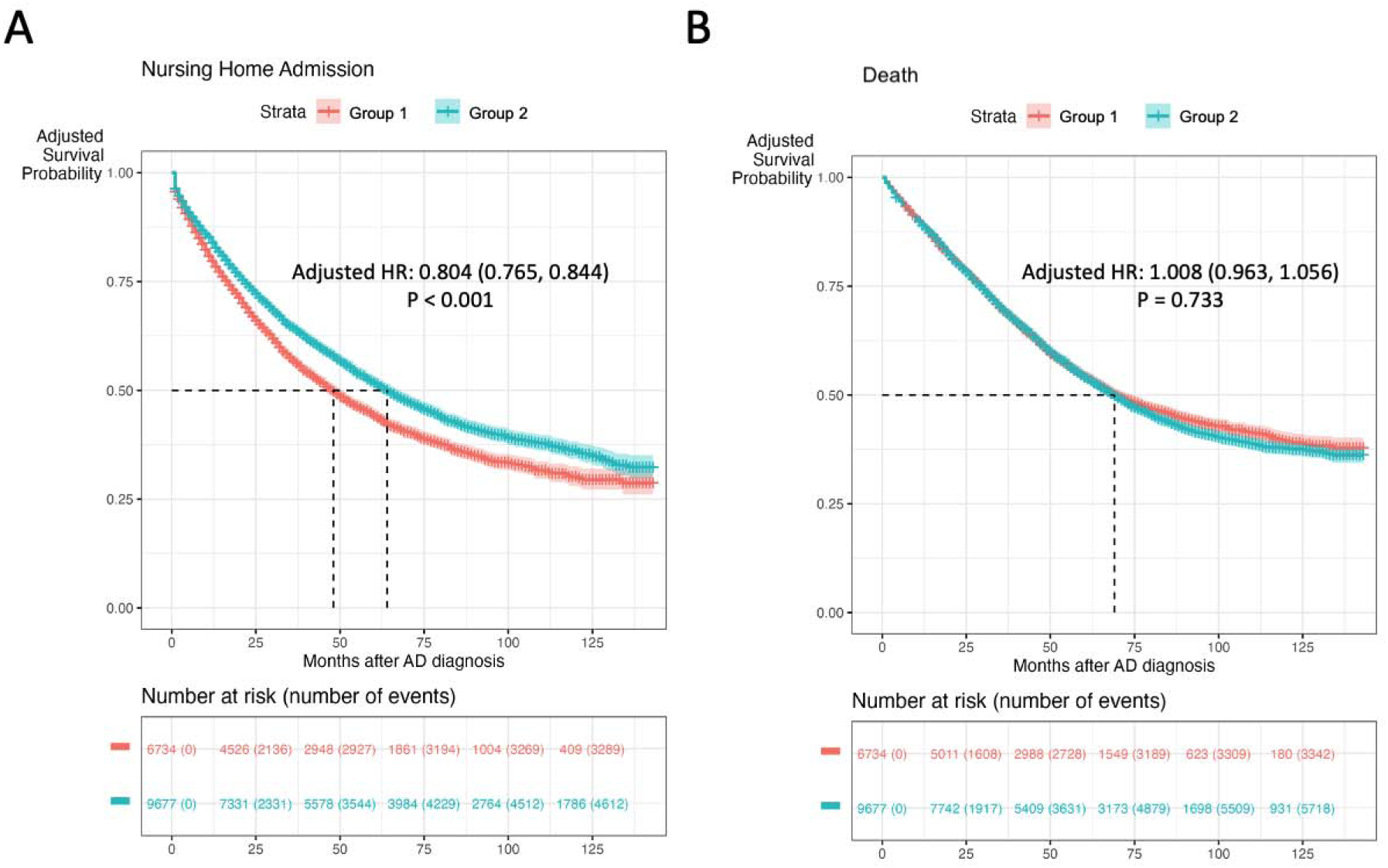
Adjusted survival curve of (A) time to nursing home and (B) time to death for AD patient groups clustered at AD diagnosis.

### Feature importance

We identified features with the largest absolute feature importance score, *i.e.,* largest differences of adjusted intensity between the groups. Features with high adjusted intensity are characteristic of a given group. Group 1 had high adjusted intensity of dementia-related diagnoses (*e.g.,* dementias, delirium dementia and amnestic and other cognitive disorders), prescriptions of AD-related medication (*e.g.,* donepezil, memantine, and rivastigmine), vascular comorbidities (*e.g.,* cerebrovascular disease, cerebrovascular disorders, cerebral ischemia, cerebral artery occlusion, cerebrovascular accident, late effects of cerebrovascular disease), neurological disorders, and epilepsy (**Figure 4A**). Group 2 had high adjusted intensity of signs and symptoms of dementia (*e.g.,* confusion, mental recall), general symptoms of decline that may indicate an increased need for care (*e.g.,* asthenia, weight decreased, malaise and fatigue, clinical decision making), factors pertaining to living situation (*e.g.,* at home, household composition, living alone), vitamin B12 / cyanocobalamin prescriptions, certain comorbidities (*e.g.,* vertigo), and medications used to treat these comorbidities (*e.g.,* amlodipine for treating hypertension, sertraline for treating depression).

**Figure 4.**
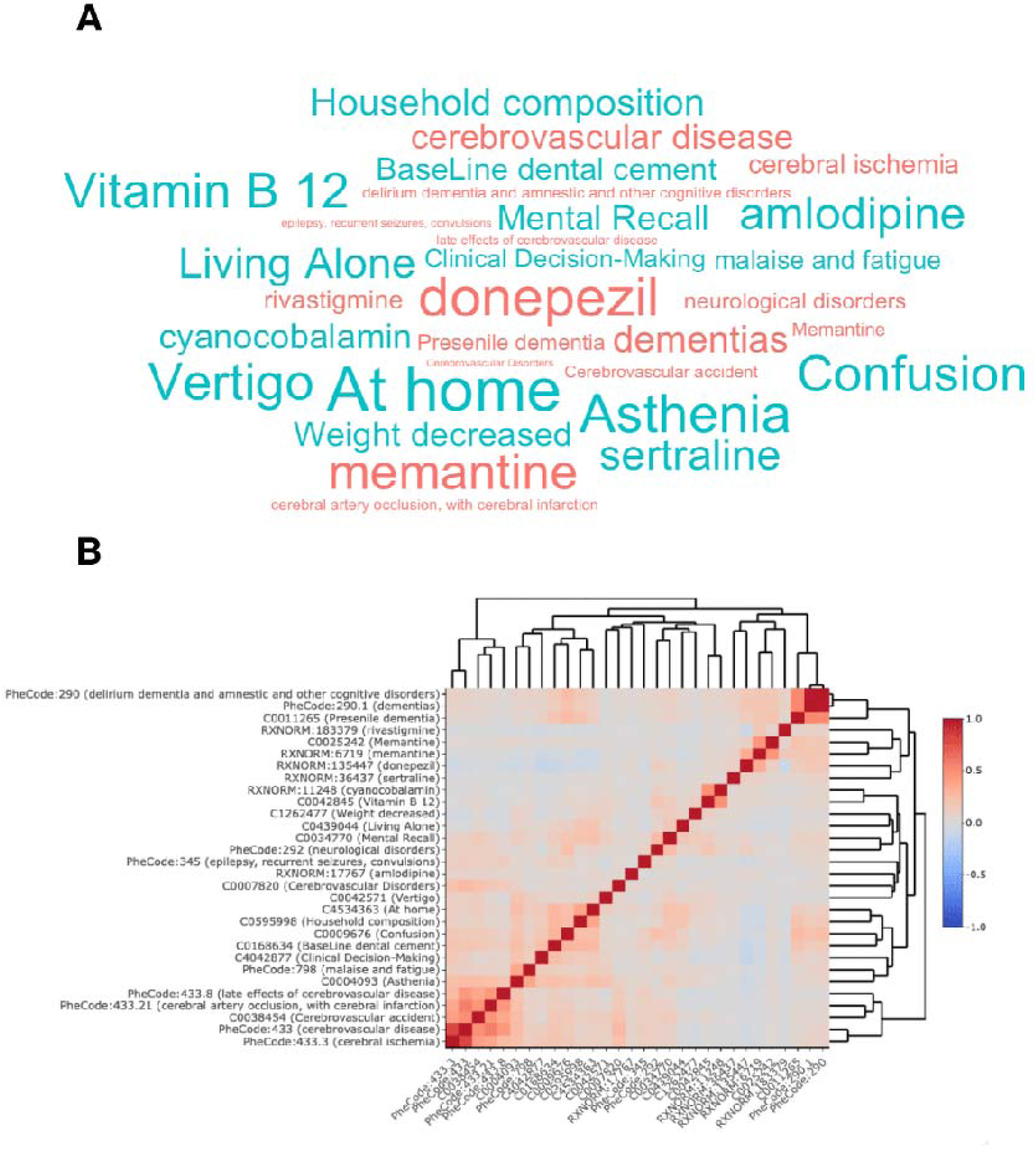
Top 15 features contributing to differences between AD patient groups clustered at AD diagnosis. Features in the color salmon were positively associated with Group 1 membership, while features in the color teal were negatively associated with Group 1 membership. (A) Word cloud of the important features. Text size indicated the relative importance of the feature. (B) Correlation matrix of the features.

We further examined the correlation between the residualized features using hierarchical clustering (**Figure 4B**). Related concepts (*e.g.,* vascular comorbidities, factors pertaining to living situation) were often correlated with each other. The features associated with each group were strongly correlated with other features in the same group.

### Prediction of outcomes one year after AD diagnosis

#### Demographic characteristics

Of the 16,411 patients in the AD cohort, 12,606 patients remained at risk for the two endpoints one year post-diagnosis (*i.e.,* alive one year post-diagnosis, not admitted to nursing home, not censored, **S-Table 8**). After updating the clustering model one year post-diagnosis, 5,759 (45.7%) patients were assigned to Group 1 and 6,847 (54.3%) to Group 2. The patients remaining at risk and included in the updated model were younger at diagnosis (age at AD diagnosis mean [SD]: Group 1: baseline: 81.4 [9.3], updated: 80.8 [9.4]; Group 2: baseline: 81.0 [8.7], updated: 80.1 [8.8]). The proportion of women remained comparable between the updated groups (updated Group 1: 3,748, 65.1%; updated Group 2: 4,413, 64.5%, *p*=.473), and the proportion of non-Hispanic White individuals was higher in the updated Group 2 (updated Group 1: 5,240, 91.0%; updated Group 2: 6,367, 93.0%; *p*<.001). The overall comorbidity burden was lower in the updated clusters in comparison to baseline (ECI mean [SD] Group 1: baseline: 11.3 [10.3], updated: 10.4 [10.0]; Group 1: baseline: 7.5 [8.6], updated: 6.4 [8.0]), but the updated Group 1 still had a higher comorbidity burden than the updated Group 2 (**S-Figure 2**).

Compared to the baseline clusters, the frequency of AD-related medication prescriptions was higher in the updated clusters one year after baseline (baseline: 77.4%; updated: 80.9%), including 45.3% preceding diagnosis and 35.6% after diagnosis (**S-Table 4**). Updated Group 1 had a higher AD-related medication prescription frequency preceding diagnosis (updated Group 1: 47.9%; updated Group 2: 43.0%; *p*<.0001) and a shorter median follow-up duration (follow up duration median [IQR]: updated Group 1: 63 [35, 86], updated Group 2: 88 [61, 126], *p*<.0001) than the updated Group 2.

### Time to AD-related outcomes

For the 12,606 patients remaining at risk one year post-diagnosis, we compared the 2-year, 5- year, and 10-year rate of nursing home admission and mortality. Without updating the cluster membership assigned at AD diagnosis, Group 1 exhibited higher rates of nursing home admission and mortality at 2-year, 5-year, and 10-year intervals than Group 2 (**S-**Table 9). After updating the cluster membership one year post-diagnosis, the updated Group 1 had a higher 2- year and 5-year rate of nursing home admission, and higher 2-year, 5-year, and 10-year mortality rate than the updated Group 2.

In survival analysis, the updated Group 2 had a lower risk of nursing home admission (HR [95% CI]=0.815 [0.766, 0.868], *p*<.001, **S-Figure 3**) than the updated Group 1, after adjusting for covariates, consistent with the baseline model. The mortality risk was again comparable between updated groups (HR [95% CI]=0.977 [0.922, 1.035], *p*=.430) after covariate adjustment.

### Feature importance

The updated Group 1 continued to exhibit a high adjusted intensity of dementia-related diagnoses and AD-related medication prescriptions (**S-Figure 4A**). Interestingly, the updated Group 1 also showed a high adjusted intensity of features associated with Parkinson disease (*e.g.,* Parkinson disease, Parkinsonian disorders, carbidopa/levodopa), which was not observed in either of the baseline groups. Further, medications used to treat benign prostatic hyperplasia (*e.g.,* tamsulosin, finasteride) and antipsychotics (*e.g.,* olanzapine) also had high adjusted intensity in the updated Group 1. In contrast, other neurological disorders and epilepsy, which were characteristic of baseline Group 1, were not prominent in the updated Group 1. Moreover, vascular comorbidities (*e.g.,* cerebrovascular disease, cerebrovascular accident), which were typical of baseline Group 1, had higher adjusted intensity in the updated Group 2. The updated Group 2 continued to be characterized by signs and symptoms of dementia, general symptoms of decline, factors pertaining to living situation, vitamin B12 / cyanocobalamin prescriptions, concomitant comorbidities, and medications used to treat these comorbidities among individuals with AD. When examining the correlation between the residualized features using hierarchical clustering, features associated with the updated groups had stronger correlations with other features in their respective groups (**S-Figure 4B**).

## DISCUSSION

This study demonstrates the feasibility of stratifying AD patients based on clinical profile available at the point of care using an unsupervised latent factor clustering model guided by knowledge-graph embeddings of EHR concepts. We stratified AD patients into two groups at AD diagnosis and crucially repeated the process one year after diagnosis for prognosticating AD-related outcomes, illustrating the potential of dynamically updating the model with more data accumulation beyond the baseline to iteratively inform prognostication. Our study has several key clinical findings. First, the patient stratification predicted one group having greater risk of future nursing home admission than the other group at AD diagnosis and again one year post-diagnosis. Second, the predicted mortality risk was comparable between the two groups stratified at AD diagnosis and one year post-diagnosis. Third, Group 1 (with higher risk for nursing home admission) was characterized by a high frequency of dementia-related diagnoses and AD-related medication prescriptions, while Group 2 was characterized by a high frequency of signs and symptoms of dementia, generalized symptoms of decline, and decreased capacity for independent living.

### Novel application of latent representation clustering for AD patient stratification

Our approach of using latent representation clustering overcomes limitations in existing approaches of using EHR data to stratify AD patients to predict AD-related outcomes. Xu et al.^25^ performed hierarchical clustering to derive AD subphenotypes for predicting the risk of AD diagnosis (but not outcomes after diagnosis) in patients identified using SNOMED-CT codes alone. Landi et al.^29^ applied deep learning to derive representations of AD patients and then hierarchical clustering to stratify AD patients into three subgroups, though the clinical utility of predicting AD-related outcomes around AD diagnosis (critical for clinical decision and prognostication discussion) was not shown. To fill this gap, He et al.^27^ identified clinically meaningful clusters predictive of mortality, by applying spectral clustering to EHR data preceding AD diagnosis, but the method relies on 40 manually selected AD-related features. Automated selection of informative features from a broad range of procedures, medications, laboratory findings, and information embedded in the clinical narratives may potentially improve the patient stratification model. Our study streamlined the process of patient stratification while improving scalability and generalizability by starting with all codified and narrative features available in the EHR and performing automated feature selection based on data-derived prior knowledge. Prior knowledge on the related features through their representations trained using separate large-scale EHR data and language models effectively harnessed existing clinical knowledge. The resulting AD patient stratification (into patient groups) based on posterior probability enabled downstream AD-related outcome prognostication.

Prediction models of AD-related outcomes that can be efficiently updated after AD diagnosis are critical for eventual clinical implementation in the real-world setting (**Figure 5**). First, diagnostic delay may influence patient stratification at AD diagnosis. Iterative updates of the prediction model after diagnosis could improve prediction of future outcomes. Second, it is unclear whether patients experience the same trajectory of decline (or variable changes in trajectory) throughout the long disease course. To systematically investigate factors that accelerate or slow the trajectory of decline, it is crucial to periodically update the prediction of AD-related outcomes. Third, with emerging disease modifying therapies such as anti-amyloid agents (*e.g.,* lecanemab, donanemab), updating the AD-related outcome prediction that incorporates AD treatment status will critically assess treatment response and clinically meaningful benefit in real-world settings.^4,5,24^

**Figure 5.**
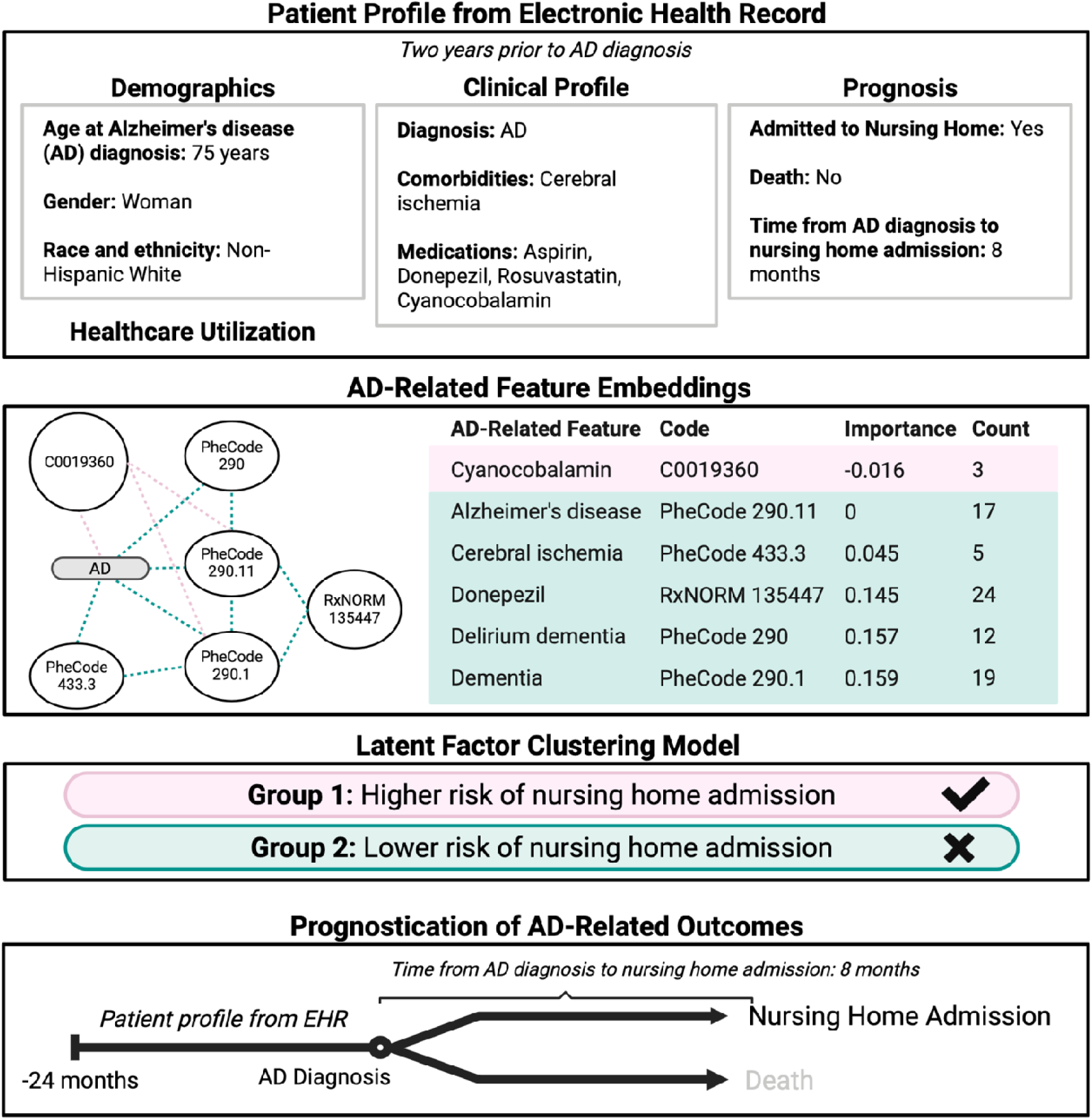
Real-world example of patient stratification. We show a hypothetical patient profile to illustrate the clinical utility of patient stratification. In the Panel entitled “Patient Profile from Electronic Health Records), the patient profile included demographics (*e.g.,* age, gender, race, ethnicity), clinical profile (*e.g.,* AD diagnosis status, comorbidities, medication prescriptions), prognosis (*e.g.,* nursing home admission status, time to death), and other relevant healthcare utilization metrics (*e.g.,* diagnosis code counts, healthcare utilization). In this example, the patient was a Non-Hispanic White woman who was diagnosed with AD at 75 years of age, treated with donepezil and cyanocobalamin. Past medical history included cerebral ischemia, managed with aspirin and rosuvastatin. The patient was admitted to a nursing home 8 months after diagnosis. In the panel entitled “AD-Related Feature Embeddings”, the patient profile further contained counts of the relevant baseline features in the two years prior to AD diagnosis (*e.g.,* AD diagnosis codes, AD-related medication prescriptions; extracted from a multi-source clinical knowledge graph) being used for prognostication and being shown for transparency. In this example, the patient had multiple counts of the diagnoses of delirium dementia (n=12), dementia (n=19), Alzheimer’s disease (n=17), and cerebral ischemia (n=5). The patient was prescribed donepezil (24 prescriptions) and cyanocobalamin (3 prescriptions). Although the patient was prescribed aspirin and rosuvastatin for cerebral ischemia, these prescription counts were not included since these are not AD-related features. (The list of AD-related features can be found in S-Table 6). In the panel entitled “Latent Factor Clustering Model”, we showed the group assignment after clustering (*e.g.,* Group 1, Group 2). In this example, the patient was assigned to Group 1 based on the baseline clinical profile, and Group 1 membership is associated with higher risk of nursing home admission. In the panel entitled “Prognostication of AD-Related Outcomes”, the patient profile included a prognostication of nursing home admission and death as competing risks. This patient was admitted to a nursing home 8 months after AD diagnosis. (Since this patient was assigned to Group 1 at AD diagnosis, she would not be reclustered one year after AD diagnosis.)

### Prognostication of the risk of nursing home admission and mortality

In survival analysis, Group 1 had a higher risk of nursing home admission (suggesting earlier need for assistance) than Group 2, when clustering patient groups at AD diagnosis and one year post-diagnosis, while the risk of mortality was comparable between the two groups when clustering at either point. Group 1 had a higher overall comorbidity burden than Group 2, with cardiometabolic diseases and other neurological disorders being the most prevalent comorbidities. Our findings are consistent with prior studies that demonstrated the association between comorbidity burden and AD disease progression.^10,41,42^ Our study sets a framework for future investigations using EHR data to address whether early effective management of common modifiable comorbidities would improve long-term cognitive and functional outcomes and reduce the risk of nursing home admission and mortality in AD patients.

### Group 1 characteristics

Dementia-related diagnoses (*e.g.,* delirium dementia and amnestic and other cognitive disorders) and prescriptions for AD-related medications to improve cognition (*e.g.,* acetylcholinesterase inhibitors)^43,44^ had high baseline feature frequencies in Group 1, when clustering at AD diagnosis and one year later. Further, AD-related medications were more frequently prescribed before diagnosis in Group 1 than Group 2. These findings suggest that patients in Group 1 likely experienced high symptom burden preceding AD diagnosis and possibly diagnostic delay (*i.e.,* diagnosed at later stages). Diagnostic delay is common among AD patients given the variable and insidious nature of the disease, with higher risk in certain demographic groups (*e.g.,* non-White individuals, lower socioeconomic status).^45,46^

Vascular comorbidities (*e.g.,* cerebrovascular disease), other neurological disorders, and epilepsy, which are known to accelerate AD decline, had high baseline frequencies in Group 1^47–53^ when clustering at AD diagnosis, but not one year later. Cerebral infarcts and vascular pathologies are independent risk factors for cognitive decline and can accelerate AD-related cognitive decline. Epilepsy often occurs in early stages of AD, and can worsen AD pathology and cognitive decline. These comorbidities, particularly when co-occurring at early AD stages, may contribute to the higher risk of nursing home admission in Group 1.

Parkinson disease and certain non-AD medications (*e.g.,* finasteride, olanzapine) had high frequency in updated Group 1, when clustering one year post-diagnosis (but not at AD diagnosis). Concomitant Parkinson disease and AD pathology can accelerate the rate of decline.^54^ While evidence supporting medications for benign prostatic hyperplasia (*e.g.,* finasteride) as disease-modifying in AD is limited,^55^ it warrants future investigation.

Antipsychotics (*e.g.,* olanzapine) are used for treating psychosis in AD^56^ as psychosis is common among AD patients and is associated with worse prognosis.^57^ While olanzapine was highlighted in our study, aripiprazole and risperidone are also used for AD psychosis.^58,59^

### Group 2 characteristics

Group 2 had a high baseline frequency of signs and symptoms of dementia (*e.g.,* confusion) when clustering at AD diagnosis and one year later. Individuals in Group 2 with signs and symptoms of dementia might be identified earlier in disease and were less likely to experience diagnostic delay than individuals in Group 1. Likewise, individuals in Group 2 had high baseline frequency of generalized symptoms of decline (*e.g.,* asthenia), suggesting decreased capacity to complete daily activities and possibly also explaining the high frequency of factors pertaining to living situations (*e.g.,* mentions of household composition) in Group 2.

Vitamin B12 and cyanocobalamin were frequently prescribed to individuals in Group 2. Randomized controlled trials and meta-analyses support vitamin B12 supplementation in improving cognitive performance in older adults with and without cognitive impairment.^60^ Although vestibular impairment may occur more frequently in AD patients than healthy older adults,^61^ the clinical significance of the high frequency of vertigo in Group 2 needs further studies. Amlodipine is a first-line antihypertensive that can reduce mortality rate but not the rate of cognitive decline in AD patients.^62^ Sertraline is a commonly used antidepressant for AD patients,^63^ while untreated depression could worsen cognition. The high frequencies of amlodipine and sertraline in Group 2 suggest treatments for common comorbidities that could otherwise worsen AD outcomes in these patients.

### Strengths and limitations

This study has several strengths. First, our latent clustering approach leveraged the rich complexity of the available EHR data, particularly the knowledge-driven feature selection of a variety of AD-related codified and narrative features, in model training. To account for similarities and relatedness among the selected features, we utilized the knowledge graph embeddings of EHR concepts to guide the projection of the observed feature counts onto a lower dimensional space, while leaving flexibility for re-weighting of each factor based on observed data. This step properly addresses the high correlations among selected features when assigning cluster membership. Second, our method is unsupervised, specifically not requiring otherwise time-consuming chart reviews. As a proof of principle, we demonstrated the feasibility to efficiently update the AD-related outcome prediction one year post-diagnosis as more clinical data become available. Third, this study uses two well-validated, generalizable and relevant clinical endpoints for AD. Nursing home admission reflects a loss of functional independence that leads to an increased need for care. Although over 40% of AD patients reside in nursing homes, nursing home admission has been underutilized as an outcome, particularly in EHR-based AD studies.^64^ While other important AD-related outcomes such as cognitive and functional measures fluctuate over time and are sparsely documented in EHR, nursing home admission and death are both concrete milestones readily ascertainable from the EHR. Finally, by prognosticating AD-related outcomes at diagnosis and then subsequently updating the prediction with more clinical data, we illustrate a pragmatic real-world clinical application. While baseline prognostication can guide early treatment initiation and tailored care management, dynamically updating the prediction model can inform more timely interventions that improve long-term outcomes.

Our study has some limitations. First, we imputed missing race and ethnicity in a small proportion of the cohort based on age and gender using a well-validated multiple imputation approach to minimize potential misclassification and retain representativeness.^37,38^ However, findings pertaining to race and ethnicity should be interpreted with caution as imputation might have introduced misclassification of race and ethnicity. Second, we clustered AD patients at AD diagnosis based on 24 months of pre-index data to strike a balance for adequate sample size and follow-up duration given the dataset contained ∼11 years of EHR data (2011-2022). Limited longitudinal pre-index data may incompletely inform baseline profile as signs and symptoms of AD precede onset by years and may inadequately capture diagnostic delay.^65,66^ Even if we had longer pre-index EHR data, there may still be a limitation of potential data incompleteness given the fragmented healthcare landscape in the US. The potential for more accurate patient stratification using longitudinal EHR data may increase with wider adoption of EHR systems and health information exchange. Third, this study did not evaluate cognitive and functional decline as endpoints. We will address this limitation in future studies when we apply novel phenotyping efforts that require minimal gold-standard labels to impute large-scale longitudinal cognitive and functional measures for AD patients. Finally, while this study included data from a single healthcare system, the AD population derived from both academic and community practices within a large catchment. When comparing to these EHR cohorts in the United States previously used in AD patient stratification studies as well as a nationwide cohort (NACC), the proportion of women in the UPMC AD cohort was similar but the proportion of racial and ethnic minorities were underrepresented (**S-Table 10**).^25–27,67^ We plan to perform external validation using independent datasets in the future.

### Future directions

Patient stratification is important for clinical management in AD.^2,68^ First, patient stratification enables systematic detection of diagnostic delay, which can inform timely diagnosis and prompt initiation of treatments that are more effective during early disease stages.^4,5^ Second, patient stratification may uncover modifiable risk factors that can be targeted to alter AD course. We could potentially use explainability tools (*e.g.,* SHapley Additive exPlanations [SHAP], Local Interpretable Model-agnostic Explanations [LIME]) to examine key risk factors that determine patient assignment to a given cluster. Clinical domain experts could further evaluate whether the risk factors as part of the feature sets of the stratification algorithm are consistent with cluster membership as sanity checks. Third, patient stratification and prognostication of AD-related outcomes can inform timely initiation of future care planning conversations with the patient and family while the patient still has decision-making capacity. Fourth, patient stratification enables the recruitment of targeted sub-populations for clinical trials and may improve trial design. Varying disease trajectory adds to the challenges of assessing treatment response for specific patient groups.^69^ There is a heightened need to identify AD patient subpopulations that may derive the greatest therapeutic benefit from novel disease-modifying therapies, given their known adverse events and high financial costs.^4,5,70^

## CONCLUSION

In summary, we applied a novel knowledge-guided unsupervised latent factor clustering approach to stratify AD patients into two groups based on baseline clinical profile at AD diagnosis. To refine the stratification, we updated the prognostication models one year post-diagnosis, aiming for a more accurate representation of the true underlying clinical trajectory. Group 1 had a higher risk of nursing home admission, though the risk of mortality was comparable between the two groups. Group 1 exhibited characteristics indicative of greater symptom burden, which may suggest diagnostic delay. In contrast, factors suggestive of gradual decline seemed more frequent in Group 2. Our study supports that patient stratification can enable outcome prognosis and possibly guide clinical management for AD patients. Future studies leveraging EHRs from multiple healthcare systems will further improve AD patient stratification.

## Data Availability

All data produced in the present study are available upon reasonable request to the authors

## ACKNOWLEDGEMENTS

This study was supported by the National Institutes of Health under award numbers R01 NS098023 and R01 NS124882 from the National Institute of General Medical Sciences. The content is solely the responsibility of the authors and does not necessarily represent the official views of the National Institutes of Health.

## AUTHOR CONTRIBUTIONS

L.W., S.Ven., M.M., M.L., R.S., S.Vis., O.L., Z.X., and T.C. contributed to the design and conceptualization of the study. L.W. and S.Ven. contributed equally as co-first authors to data analysis and manuscript writing. M.M., R.S., S.Vis., O.L., Z.X., and T.C. contributed to data acquisition and manuscript writing. Z.X. and T.C. contributed equally as co-senior authors and jointly supervised this work. All authors have reviewed and approved the final manuscript.

