## Supplemental Materials for "Stratification of Alzheimer’s Disease Patients Using Knowledge-Guided Unsupervised Latent Factor Clustering with Electronic Health Record Data"

### SUPPLEMENTARY METHODS

To ensure the scalability of our model training, we implemented an algorithm based on variational Gaussian approximation and Expectation-Maximization [1]. At model convergence, the posterior probability of membership of the latent classes was calculated for each patient. The patient was assigned exclusively to the class with the highest posterior probability. The details are outlined below:

The complete-data log likelihood is

$$\log p(\boldsymbol{X},\boldsymbol{W},\boldsymbol{Y};\boldsymbol{B},\boldsymbol{\Lambda},a_0,a_1) = \sum_{i=1}^{N}\{\log p(\boldsymbol{X}_i \mid \boldsymbol{W}_i,U_i,Y_i;\boldsymbol{B},\boldsymbol{\Lambda},\pi_i) + \log p(\boldsymbol{W}_i) + \log p(Y_i;\pi_i)\}$$

Since $\boldsymbol{W}$ and $\boldsymbol{Y}$ are latent, the evaluations of the log-likelihood of the observed data

$$\ell_L(\boldsymbol{B},\boldsymbol{\Lambda},a_0,a_1) = \log p(\boldsymbol{X};\boldsymbol{B},\boldsymbol{\Lambda},a_0,a_1) = \log \int\int p(\boldsymbol{X},\boldsymbol{W},\boldsymbol{Y};\boldsymbol{B},\boldsymbol{\Lambda},a_0,a_1)d\boldsymbol{W}d\boldsymbol{Y}$$

is intractable, as well as its maximization with respect to $(\boldsymbol{B},\boldsymbol{\Lambda})$.

Variational approximation presents an alternative parameter estimation framework by using a computationally convenient approximating density in place of a more complex posterior density [2]. Using computationally convenient Gaussian densities, complex posterior distributions are approximated by minimizing the Kullback-Leibler (KL) divergence between the true and the approximating densities and therefore, reducing the computational overhead. Here instead of directly maximizing the log likelihood, we maximize the evidence lower bound (ELBO):

$$\begin{aligned}\mathscr{O}(\boldsymbol{B},\boldsymbol{\Lambda},q) &= \log p(\boldsymbol{X} \mid \boldsymbol{U};\boldsymbol{B},\boldsymbol{\Lambda}) - KL[q(\boldsymbol{W},\boldsymbol{Y} \mid \boldsymbol{X},\boldsymbol{U}), p(\boldsymbol{W},\boldsymbol{Y} \mid \boldsymbol{X},\boldsymbol{U};\boldsymbol{B},\boldsymbol{\Lambda}))]\\ &= \mathbb{E}_q[\log p(\boldsymbol{X},\boldsymbol{W},\boldsymbol{Y} \mid \boldsymbol{U};\boldsymbol{B},\boldsymbol{\Lambda})] - \mathbb{E}_q[\log q(\boldsymbol{W},\boldsymbol{Y} \mid \boldsymbol{X},\boldsymbol{U})]\end{aligned}$$

We used a mixture Gaussian approximation for

$$q(\boldsymbol{W}_i,Y_i=y \mid \boldsymbol{X}_i,U_i) = \underbrace{q(\boldsymbol{W}_i \mid Y_i=y,\boldsymbol{X}_i,U_i)}_{q_y(\boldsymbol{W}_i)}\underbrace{P(Y_i=y \mid \boldsymbol{X}_i,U_i)}_{\gamma_i}$$

with

$$q_y(\boldsymbol{W}_i \mid Y_i=y,\boldsymbol{X}_i,U_i) \sim \mathscr{N}(\boldsymbol{m}_i^{(y)},\boldsymbol{s}_i^{(y)}) \;\; for \;\; y=0,1.$$

Let $\boldsymbol{\Theta} = \{\boldsymbol{B}, \boldsymbol{\Lambda}, a_0, a_1\}$ and $\boldsymbol{\Omega} = (\boldsymbol{M}^{(0)}, \boldsymbol{S}^{(0)}, \boldsymbol{M}^{(1)}, \boldsymbol{S}^{(1)})$, our objective function is

$$
\begin{aligned}
\mathscr{O}(\boldsymbol{\Theta}, \boldsymbol{\Omega}; \boldsymbol{X}, \boldsymbol{U}) = \sum_{y=0}^{1} \mathbf{1}_n^{\mathsf{T}} &\left[\widehat{\boldsymbol{\Gamma}}_p^y (\mathbf{1} - \widehat{\boldsymbol{\Gamma}}_p)^{1-y} \odot \left(\boldsymbol{X} \odot [(\boldsymbol{D}^{(y)}\boldsymbol{B} + \boldsymbol{M}^{(y)})\boldsymbol{\Lambda}^{1/2}\boldsymbol{V}^{\mathsf{T}}] - \mathbb{E}_{q_y}\left\{\exp[(\boldsymbol{D}^{(y)}\boldsymbol{B} + \boldsymbol{W})\boldsymbol{\Lambda}^{1/2}\boldsymbol{V}^{\mathsf{T}}]\right\}\right)\right] \mathbf{1}_p \\
&- \frac{1}{2}\mathbf{1}_n^{\mathsf{T}} \left\{\widehat{\boldsymbol{\Gamma}}_q^y (\mathbf{1} - \widehat{\boldsymbol{\Gamma}}_q)^{1-y} \odot (\boldsymbol{M}^{(y)} \odot \boldsymbol{M}^{(y)} + \boldsymbol{S}^{(y)} \odot \boldsymbol{S}^{(y)} - 2\log(\boldsymbol{S}^{(y)}) - \mathbf{1}_{n\times q})\right\} \mathbf{1}_q \\
&+ \widehat{\boldsymbol{\Gamma}}^y (\mathbf{1} - \widehat{\boldsymbol{\Gamma}})^{1-y} \left[y \log\{expit(a_0 + a_1\boldsymbol{U})\} + (1-y)\log\{1 - expit(a_0 + a_1\boldsymbol{U})\}\right] \\
&- \widehat{\boldsymbol{\Gamma}}^y (\mathbf{1} - \widehat{\boldsymbol{\Gamma}})^{1-y} \left[\log\{\widehat{\boldsymbol{\Gamma}}^y (1 - \widehat{\boldsymbol{\Gamma}}^y)\}\right]
\end{aligned}
$$

where $\boldsymbol{\Gamma} = (\gamma_1, \cdots, \gamma_N) \in \mathbb{R}^N$.

The algorithm alternates between two steps until convergence:

- **E-Step:** Let $\widehat{\mathscr{J}_i^{(y)}} = \mathscr{J}(\widehat{\boldsymbol{\Theta}}, \widehat{\boldsymbol{\Omega}}; \boldsymbol{X}_i, \boldsymbol{D}_i^{(y)}) = \mathbb{E}_{q_y}[\log p(\boldsymbol{X}_i, \boldsymbol{W}_i, Y_i = y \mid U_i)] - \mathbb{E}_{q_y}[\log q_y(\boldsymbol{W}_i \mid \boldsymbol{X}_i, Y_i = y, U_i)]$, we update
$$\widehat{\gamma}_i = \frac{\exp(\widehat{\mathscr{J}_i^{(1)}})}{\exp(\widehat{\mathscr{J}_i^{(1)}}) + \exp(\widehat{\mathscr{J}_i^{(0)}})}$$

- **M-Step:** Optimize for parameters $(\widehat{\boldsymbol{\Theta}}, \widehat{\boldsymbol{\Omega}}) = \arg\max_{\boldsymbol{\Theta},\boldsymbol{\Omega}} \mathscr{O}(\boldsymbol{\Theta}, \boldsymbol{\Omega}; \boldsymbol{X}, \boldsymbol{U})$, by alternating between the following two steps until convergence:

  - **Optimizing for $\boldsymbol{B}$, $\boldsymbol{\Lambda}$, $\boldsymbol{\Omega}$:** We first optimize for these parameters using the globally convergent method of moving asymptotes (MMA) algorithm for gradient-based local optimization implemented in the `NLOPT` optimization library in `C++` [3,4]. We use box constraints for the positive model parameters $\boldsymbol{\Lambda}$ and the variational parameters $\boldsymbol{S}$. The blockwise gradients are derived as follows:

$$
\begin{aligned}
\frac{\partial \mathscr{O}}{\partial \boldsymbol{M}^{(y)}} &= \left\{\boldsymbol{\Gamma}^y (\mathbf{1} - \boldsymbol{\Gamma})^{1-y} \mathbf{1}_p^{\mathsf{T}} \odot (\boldsymbol{X} - \boldsymbol{A}^{(y)})\right\} \boldsymbol{V}\boldsymbol{\Lambda}^{1/2} - \boldsymbol{M} \\
\frac{\partial \mathscr{O}}{\partial \boldsymbol{S}^{(y)}} &= \boldsymbol{S}^{\oslash} - \boldsymbol{S} - \left\{\boldsymbol{\Gamma}^y (\mathbf{1} - \boldsymbol{\Gamma})^{1-y} \mathbf{1}_p^{\mathsf{T}} \odot \boldsymbol{A}^{(y)}\right\} (\boldsymbol{V}\boldsymbol{\Lambda}^{1/2} \odot \boldsymbol{V}\boldsymbol{\Lambda}^{1/2}) \odot \boldsymbol{S} \\
\frac{\partial \mathscr{O}}{\partial \boldsymbol{B}} &= \boldsymbol{\Lambda}^{1/2}\boldsymbol{V}\left\{(\boldsymbol{X} - \boldsymbol{A}^{(0)})^{\mathsf{T}} (1 - \boldsymbol{\Gamma}) \mathbf{1}_d^{\mathsf{T}} \odot \boldsymbol{D}^{(0)} + (\boldsymbol{X} - \boldsymbol{A}^{(1)})^{\mathsf{T}} \boldsymbol{\Gamma} \mathbf{1}_d^{\mathsf{T}} \odot \boldsymbol{D}^{(1)}\right\} \\
\frac{\partial \mathscr{O}}{\partial \boldsymbol{\Lambda}^{1/2}} &= diag\Bigg\{\boldsymbol{V}^{\mathsf{T}}\Bigg[(\boldsymbol{X} - \left\{(\mathbf{1} - \boldsymbol{\Gamma})\mathbf{1}_p^{\mathsf{T}} \odot \boldsymbol{A}^{(0)} + \boldsymbol{\Gamma}\mathbf{1}_p^{\mathsf{T}} \odot \boldsymbol{A}^{(1)}\right\})^{\mathsf{T}} (\boldsymbol{D}_L\boldsymbol{B} + \boldsymbol{M}_L) - \\
&\qquad [\left\{(\mathbf{1} - \boldsymbol{\Gamma})\mathbf{1}_p^{\mathsf{T}} \odot \boldsymbol{A}^{(0)} + \boldsymbol{\Gamma}\mathbf{1}_p^{\mathsf{T}} \odot \boldsymbol{A}^{(1)}\right\}^{\mathsf{T}} (\boldsymbol{S} \odot \boldsymbol{S})] \odot \boldsymbol{V}\boldsymbol{\Lambda}^{1/2}\Bigg]\Bigg\}
\end{aligned}
$$

– **Optimizing for $a_0$ and $a_1$:** We then update $(a_0, a_1)$ by optimizing the objective function

$$(\widehat{a}_0, \widehat{a}_1) = \arg\max_{a_0, a_1} \sum_{i=1}^{N} \widehat{\gamma}_i \log\{expit(a_0 + a_1 U_i)\} + (1 - \widehat{\gamma}_i) \log\{1 - expit(a_0 + a_1 U_i)\}$$

We can do this by fitting weighted logistic regression with outcomes $(\mathbf{0}_N, \mathbf{1}_N)$, weights $(\mathbf{1} - \widehat{\boldsymbol{\gamma}}, \widehat{\boldsymbol{\gamma}})$ and covariates $(\boldsymbol{U}, \boldsymbol{U})$.

4. Steven G. Johnson, The NLopt nonlinear-optimization package, http://github.com/stevengj/nlopt.

### SUPPLEMENTARY ANALYSES

#### Comparison to Benchmark Models

We compared the results of the Poisson-LogNormal Mixture Model (PLMM) clustering method at AD diagnosis with two commonly used benchmark models, K-means clustering and Gaussian Mixture Model. To fit these models, we first performed logarithmic transformation of baseline EHR feature counts (*i.e.,* x -> log[x+1]) and used principal component analysis to reduce the dimensionality of the feature space. We then examined the association between cluster membership at diagnosis and AD-related outcomes in competing risk Cox proportional hazard models. Compared to the PLMM model, K-means and Gaussian mixture modeling produced clusters that were less distinct in terms of their risk of nursing home admission (HR [95% CI]: K-means=0.954 [0.902, 1.014] *p*=.07; Gaussian Mixture Model=0.951 [0.899, 1.010], *p*=.07; PLMM=0.804 [0.765, 0.844], *p*<.001, **S-Figure 6**). This demonstrates the utility of the PLMM method in terms of stratifying AD patients into disparate risk groups.

#### Sensitivity Analysis Using Different Cluster Counts

We calculated the approximate Bayesian Information Criterion (BIC), which balances model fit and complexity, for models with K=2, 3, 4, and 5 clusters. Lower BIC values indicate better model fit. K=2 yielded the lowest BIC value, suggesting that two clusters provided the best balance between fit and simplicity (**S-Table 2**). Therefore, we selected the model with 2 clusters. In a sensitivity analysis assessing the time to nursing home admission and time to death for K=3, Group 2 and Group 3 exhibited similar risks for both nursing home admission and death, supporting the decision for using two clusters, which had adequately captured the patient heterogeneity (**S-Figure 5**).

**Stratified Analysis Based on Baseline Healthcare Utilization**

To assess the robustness of the clustering approach given the potential confounding due to healthcare utilization differences, we conducted a stratified analysis by dividing the cohort into low- vs high- healthcare utilization subgroups based on total number of ICD codes plus encounter notes (e.g., below and above or equal to the median). We then performed clustering separately within each healthcare utilization subgroup. We evaluated the stability of cluster assignments using Adjusted Rand index (ARI) metrics, which is 0.65 for the high utilization group and 0.72 for the low utilization group (S-Table 11, S-Table 12). ARI is a measure of similarity between clusters, with 0 indicating no similarity and 1 indicating complete agreement. This finding supported the stability of the clustering approach despite differences in healthcare utilization levels.

### SUPPLEMENTARY TABLES

**S-Table 1.** AD data mart specifications.

| Code | Description |
|---|---|
| ICD-9 290.0 | Senile dementia, uncomplicated |
| ICD-9 290.1 | Presenile dementia |
| ICD-9 290.10 | Presenile dementia, uncomplicated |
| ICD-9 290.11 | Presenile dementia with delirium |
| ICD-9 290.12 | Presenile dementia with delusional features |
| ICD-9 290.13 | Presenile dementia with depressive features |
| ICD-9 290.20 | Senile dementia with delusional features |
| ICD-9 290.21 | Senile dementia with depressive features |
| ICD-9 290.3 | Senile dementia with delirium |
| ICD-9 294.20 | Dementia, unspecified, without behavioral disturbance |
| ICD-9 294.21 | Dementia, unspecified, with behavioral disturbance |
| ICD-9 331.0 | Alzheimer’s disease |
| ICD-10 F03.90 | Unspecified dementia without behavioral disturbance |
| ICD-10 F03.91 | Unspecified dementia with behavioral disturbance |
| ICD-10 G30 | Alzheimer's disease |
| ICD-10 G30.0 | Alzheimer's disease with early onset |
| ICD-10 G30.1 | Alzheimer's disease with late onset |
| ICD-10 G30.8 | Other Alzheimer's disease |
| ICD-10 G30.9 | Alzheimer's disease, unspecified |

Note: Patients were included in the data mart if they had at least one of the AD related diagnosis codes. The data mart is the starting point to more accurately identify AD patients.

**S-Table 2.** Bayesian Information Criterion (BIC) values for varying number of clusters (K).

| | K=2 | K=3 | K=4 | K=5 |
|---|---|---|---|---|
| Approximate BIC | -1183.51 | -1179.63 | -1150.01 | -1100.94 |

Note: BIC, Bayesian Information Criterion, with lower BIC value indicating better model fit; K, number of clusters

**S-Table 3.** List of nursing home admission codes.

| Code | Description |
|---|---|
| **Assisted living** | |
| CPT 99321 | Domiciliary or rest home visit for the evaluation and management of a new patient; the presenting problems are of low severity |
| CPT 99322 | Domiciliary or rest home visit for the evaluation and management of a new patient; the presenting problems are of moderate severity |
| CPT 99323 | Domiciliary or rest home visit for the evaluation and management of a new patient; the presenting problems are of high complexity |
| CPT 99331 | Domiciliary or rest home visit for the evaluation and management of a new patient; the patient is stable, recovering or improving |
| CPT 99332 | Domiciliary or rest home visit for the evaluation and management of a new patient; the patient is responding inadequately to therapy or has developed a minor complication |
| CPT 99333 | Domiciliary or rest home visit for the evaluation and management of a new patient; the patient is unstable or has developed a significant complication or a significant new problem |
| ICD-9 V60.89 | Other specified housing or economic circumstances |
| ICD-10 Z74 | Problems related to care provider dependency |
| ICD-9 V63.2 | Person awaiting admission to adequate facility elsewhere |
| ICD-10 Z75.1 | Person awaiting admission to adequate facility elsewhere |
| **Long-term care** | |
| CPT 99301 | Evaluation and management of a new or established patient involving an annual nursing facility assessment: 30 minutes at the bedside |
| CPT 99302 | Evaluation and management of a new or established patient involving an annual nursing facility assessment of a complication or a new problem: 40 minutes at the bedside |
| CPT 99303 | Evaluation and management of a new or established patient involving an annual nursing facility assessment at the time of initial admission to the facility: 50 minutes at the bedside |
| CPT 99304 | Initial nursing facility care 25 min |
| CPT 99305 | Initial nursing facility care 35 min |
| CPT 99306 | Initial nursing facility care 45 min |
| CPT 99307 | Subsequent nursing facility care, per day, for the evaluation and management of a patient: 10 minutes at the bedside |
| CPT 99308 | Subsequent nursing facility care, per day, for the evaluation and management of a patient: 15 minutes at the bedside |
| CPT 99309 | Subsequent nursing facility care, per day, for the evaluation and management of a patient: 25 minutes at the bedside |
| CPT 99310 | Subsequent nursing facility care, per day, for the evaluation and management of a patient: 35 minutes at the bedside |
| CPT 99311 | Subsequent nursing facility care, per day, for the evaluation and management of a new or established patient: 15 minutes at the bedside |
| CPT 99312 | Subsequent nursing facility care, per day, for the evaluation and management of a new or established patient who is responding inadequately to therapy or has developed a minor complication: 25 minutes at the bedside |

| | |
|---|---|
| CPT 99313 | Subsequent nursing facility care, per day, for the evaluation and management of a new or established patient who has developed a significant complication or a new problem: 35 minutes at the bedside |
| CPT 99315 | Nursing facility discharge day management; 30 minutes or less |
| CPT 99316 | Nursing facility discharge day management; more than 30 minutes |
| CPT 99318 | Evaluation and management of a patient involving an annual nursing facility assessment |
| CPT 99379 | Physician supervision of a nursing facility patient (patient not present) requiring complex and multidisciplinary care; 15–29 minutes |
| CPT 99380 | Physician supervision of a nursing facility patient (patient not present) requiring complex and multidisciplinary care modalities; 30 minutes or more |
| CPT 99199 | Unlisted special service, procedure, or report |
| ICD-9 V60.6 | Person living in residential institution |
| ICD-10 Z59.3 | Problems related to living in residential institution |
| ICD-9 E849.7 | Accidents occurring in residential institution |
| ICD-10 Y92.12 | Nursing home as the place of occurrence of the external cause |
| ICD-9 E849.0 | Accidents occurring in hospital |
| ICD-10 Y92.23 | Hospital as the place of occurrence of the external cause |
| ICD-9 V70.3 | Other general medical examination for administrative purposes |
| ICD-10 Z02.2 | Encounter for examination for admission to residential institution |
| HCPCS G0066 | Physician supervision of a nursing facility patient (patient not present); 30 minutes or more per month |
| HCPCS G9685 | Physician service or other qualified health care professional for the evaluation and management of a beneficiary's acute change in condition in a nursing facility |
| **End-of-life care** | |
| CPT 99377 | Under Care Plan Oversight Services. Report this service when the provider supervises and coordinates the care provided to a hospice patient. |
| CPT 99378 | Under Care Plan Oversight Services. Report this service when the provider supervises and coordinates the care provided to a hospice patient. |
| ICD-9 E876.8 | Other specified misadventures during medical care |
| ICD-10 Y66 | Non-administration of surgical and medical care |
| ICD-9 V65.49 | Other specified counseling |
| ICD-9 V65.8 | Other reasons for seeking consultation |
| ICD-10 Z76.89 | Persons encountering health services in other specified circumstances |
| ICD-9 V66.7 | Encounter for palliative care |
| ICD-10 Z51.5 | Encounter for palliative care |
| HCPCS G0299 | Direct skilled nursing services of a registered nurse (RN) in the home health or hospice setting, each 15 minutes |
| HCPCS G0300 | Direct skilled nursing services of a licensed practical nurse (LPN) in the home health or hospice setting, each 15 minutes |
| HCPCS Q5001 | Hospice or home health care provided in patient's home/residence |
| HCPCS Q5002 | Hospice or home health care provided in assisted living facility |
| HCPCS Q5003 | Hospice care provided in nursing long term care facility (LTC) or non-skilled nursing facility (NF) |
| HCPCS Q5004 | Hospice care provided in skilled nursing facility (SNF) |
| HCPCS Q5005 | Hospice care provided in inpatient hospital |
| HCPCS Q5006 | Hospice care provided in inpatient hospice facility |
| HCPCS Q5007 | Hospice care provided in long term care facility |

| | |
|---|---|
| HCPCS Q5008 | Hospice care provided in inpatient psychiatric facility |
| HCPCS Q5009 | Hospice or home health care provided in place not otherwise specified (NOS) |
| HCPCS Q5010 | Hospice home care provided in a hospice facility |
| HCPCS T2042 | Hospice routine home care per diem |
| HCPCS T2043 | Hospice continuous home care per hour |
| HCPCS T2044 | Hospice inpatient respite care per diem |
| HCPCS T2045 | Hospice general inpatient care per diem |
| HCPCS T2046 | Hospice long term care, room and board only per diem |

**S-Table 4.** List of AD-related medications.

| Brand Name | Generic Name | Treatment Class |
|---|---|---|
| Adlarity | Donepezil | Acetylcholinesterase Inhibitor |
| Aricept | Donepezil | Acetylcholinesterase Inhibitor |
| Exelon | Rivastigmine | Acetylcholinesterase Inhibitor |
| Razadyne ER | Galantamine | Acetylcholinesterase Inhibitor |
| Razadyne [DSC] | Galantamine | Acetylcholinesterase Inhibitor |
| Namenda | Memantine | N-Methyl-D-Aspartate (NMDA) Receptor Antagonist |
| Namenda Titration Pak | Memantine | N-Methyl-D-Aspartate (NMDA) Receptor Antagonist |
| Namenda XR | Memantine | N-Methyl-D-Aspartate (NMDA) Receptor Antagonist |
| Namenda XR Titration Pack [DSC] | Memantine | N-Methyl-D-Aspartate (NMDA) Receptor Antagonist |
| Aduhelm | Aducanumab | Anti-Amyloid Monoclonal Antibody |
| Leqembi | Lecanemamb-irmb | Anti-Amyloid Monoclonal Antibody |
| Namzaric | Donepezil and memantine | Acetylcholinesterase Inhibitor; N-Methyl-D-Aspartate (NMDA) Receptor Antagonist |

**S-Table 5.** Importance and prevalence of AD-related features included in the clustering model at AD diagnosis.

| Feature | Description | Feature Importance | Prevalence | | |
|---|---|---|---|---|---|
| | | | Overall | Group 1 | Group 2 |
| RXNORM:135447 | Donepezil | 0.1453 | 42.57% | 45.28% | 40.06% |
| PheCode:433 | Cerebrovascular disease | 0.1098 | 36.31% | 40.07% | 32.82% |
| RXNORM:6719 | Memantine | 0.0979 | 20.09% | 18.34% | 21.71% |
| PheCode:290.1 | Dementias | 0.0518 | 100.00% | 100.00% | 100.00% |
| PheCode:290 | Delirium dementia and amnestic and other cognitive disorders | 0.0501 | 100.00% | 100.00% | 100.00% |
| PheCode:433.3 | Cerebral ischemia | 0.0446 | 27.87% | 31.75% | 24.27% |
| PheCode:292 | Neurological disorders | 0.0365 | 52.63% | 62.22% | 43.76% |
| PheCode:433.21 | Cerebral artery occlusion, with cerebral infarction | 0.0221 | 8.44% | 9.34% | 7.60% |
| C0011265 | Presenile dementia | 0.0158 | 66.44% | 76.37% | 57.25% |
| RXNORM:183379 | Rivastigmine | 0.0141 | 6.70% | 6.68% | 6.71% |
| C0038454 | Cerebrovascular accident | 0.0079 | 26.17% | 35.39% | 17.63% |
| PheCode:433.8 | Late effects of cerebrovascular disease | 0.0069 | 4.10% | 4.57% | 3.66% |
| PheCode:345 | Epilepsy, recurrent seizures, convulsions | 0.0060 | 6.78% | 7.78% | 5.86% |
| C0025242 | Memantine | 0.0058 | 8.69% | 9.86% | 7.60% |
| C0007820 | Cerebrovascular Disorders | 0.0048 | 3.87% | 4.95% | 2.87% |
| C0007787 | Transient Ischemic Attack | 0.0046 | 7.85% | 10.48% | 5.42% |
| C0002395 | Alzheimer's Disease | 0.0045 | 54.34% | 61.39% | 47.81% |
| PheCode:332 | Parkinson's disease | 0.0043 | 4.86% | 5.30% | 4.44% |
| RXNORM:114477 | Levetiracetam | 0.0033 | 3.01% | 3.61% | 2.46% |
| C0527316 | Donepezil | 0.0031 | 16.98% | 21.19% | 13.08% |
| PheCode:430 | Intracranial hemorrhage | 0.0028 | 2.57% | 2.78% | 2.38% |
| C0003537 | Aphasia | 0.0015 | 4.26% | 5.55% | 3.07% |
| C0011269 | Dementia, Vascular | 0.0013 | 3.65% | 4.81% | 2.57% |
| PheCode:818 | Intracranial hemorrhage (injury) | 0.0010 | 1.80% | 1.84% | 1.76% |
| PheCode:430.2 | Intracerebral hemorrhage | 0.0006 | 1.09% | 1.31% | 0.89% |
| C0242422 | Parkinsonian Disorders | 0.0004 | 2.68% | 3.64% | 1.80% |
| RXNORM:103990 | Carbidopa/levodopa | 0.0002 | 2.81% | 3.53% | 2.13% |
| C0085220 | Cerebral Amyloid Angiopathy | 0.0001 | 0.19% | 0.21% | 0.18% |
| C0751587 | CADASIL Syndrome | 0.0000 | 0.06% | 0.06% | 0.07% |

| | | | | | |
|---|---|---|---|---|---|
| C3662068 | Static encephalopathy | 0.0000 | 0.04% | 0.06% | 0.03% |
| C0016957 | Galactosylceramidase | 0.0000 | 0.00% | 0.00% | 0.00% |
| C0393483 | Brainstem encephalitis | 0.0000 | 0.00% | 0.00% | 0.00% |
| C4763868 | Cocaine use disorder | 0.0000 | 0.01% | 0.03% | 0.00% |
| PheCode:290.11 | Alzheimer's disease | 0.0000 | 99.94% | 99.91% | 99.96% |
| C0260942 | Encounter due to screening for depression | 0.0000 | 0.09% | 0.15% | 0.04% |
| RXNORM:1791685 | Pimavanserin | 0.0000 | 0.04% | 0.06% | 0.01% |
| PheCode:333.3 | Tics and choreas | 0.0000 | 0.06% | 0.12% | 0.00% |
| C0030567 | Parkinson disease | -0.0001 | 5.59% | 7.26% | 4.04% |
| RXNORM:4637 | Galantamine | -0.0001 | 0.69% | 0.73% | 0.65% |
| C0041296 | Tuberculosis | -0.0003 | 0.91% | 1.41% | 0.44% |
| C3714756 | Intellectual disability | -0.0009 | 0.90% | 1.32% | 0.51% |
| C0011127 | Pressure ulcer | -0.0011 | 2.13% | 3.40% | 0.95% |
| C0123091 | Quetiapine | -0.0018 | 2.05% | 3.50% | 0.70% |
| C4484264 | Clark | -0.0018 | 2.03% | 3.16% | 0.99% |
| C0022983 | Laminectomy | -0.0025 | 2.96% | 4.60% | 1.43% |
| RXNORM:61381 | Olanzapine | -0.0034 | 2.81% | 4.34% | 1.39% |
| RXNORM:25025 | Finasteride | -0.0042 | 3.52% | 4.71% | 2.42% |
| PheCode:292.4 | Altered mental status | -0.0043 | 23.53% | 29.91% | 17.63% |
| C0518460 | Bathing self care | -0.0052 | 6.33% | 10.05% | 2.87% |
| C0008845 | Citalopram | -0.0053 | 4.78% | 7.44% | 2.31% |
| C0278061 | Abnormal mental state | -0.0075 | 14.65% | 20.52% | 9.21% |
| RXNORM:43611 | Latanoprost | -0.0090 | 3.14% | 4.89% | 1.53% |
| C0019360 | Herpes zoster disease | -0.0094 | 8.64% | 13.38% | 4.25% |
| C0011168 | Deglutition disorders | -0.0095 | 7.94% | 12.67% | 3.56% |
| RXNORM:35636 | Risperidone | -0.0098 | 3.88% | 5.90% | 2.01% |
| PheCode:295 | Schizophrenia and other psychotic disorders | -0.0104 | 9.47% | 12.68% | 6.50% |
| C0428977 | Bradycardia | -0.0110 | 8.52% | 13.60% | 3.81% |
| RXNORM:221147 | Polyethylene glycol 3350 | -0.0116 | 10.02% | 16.29% | 4.21% |
| C0430533 | Dental diagnostic procedure | -0.0132 | 13.22% | 20.58% | 6.39% |
| PheCode:350.2 | Abnormality of gait | -0.0137 | 15.24% | 21.01% | 9.89% |
| RXNORM:321988 | Escitalopram | -0.0209 | 6.20% | 9.10% | 3.51% |
| C0034991 | Rehabilitation therapy | -0.0217 | 14.14% | 23.60% | 5.39% |
| RXNORM:51272 | Quetiapine | -0.0225 | 5.87% | 9.24% | 2.75% |
| RXNORM:77492 | Tamsulosin | -0.0226 | 8.76% | 12.73% | 5.09% |
| RXNORM:15996 | Mirtazapine | -0.0237 | 5.50% | 9.03% | 2.24% |
| RXNORM:10737 | Trazodone | -0.0259 | 5.75% | 9.33% | 2.45% |
| PheCode:591 | Urinary tract infection | -0.0295 | 23.25% | 30.56% | 16.49% |
| C0011053 | Deafness | -0.0306 | 14.89% | 23.43% | 6.97% |
| C0757844 | TNFSF13 protein, human | -0.0319 | 20.75% | 32.76% | 9.63% |
| C0206275 | Widowhood | -0.0325 | 11.70% | 19.02% | 4.92% |

| | | | | | |
|---|---|---|---|---|---|
| C3809991 | Immunodeficiency, common variable | -0.0336 | 17.84% | 27.58% | 8.81% |
| C0042029 | Urinary tract infection | -0.0364 | 17.59% | 27.43% | 8.47% |
| C1856053 | Hydranencephaly with renal aplasia-dysplasia | -0.0375 | 21.25% | 33.75% | 9.67% |
| PheCode:798 | Malaise and fatigue | -0.0447 | 29.07% | 41.27% | 17.78% |
| C4042877 | Clinical decision-making | -0.0521 | 42.17% | 57.71% | 27.79% |
| C0034770 | Mental recall | -0.0553 | 34.61% | 51.40% | 19.06% |
| RXNORM:36437 | Sertraline | -0.0576 | 10.80% | 16.05% | 5.94% |
| RXNORM:11248 | Cyanocobalamin | -0.0577 | 14.48% | 22.82% | 6.75% |
| C0042571 | Vertigo | -0.0638 | 26.54% | 41.00% | 13.15% |
| C0439044 | Living alone | -0.0642 | 20.13% | 33.41% | 7.82% |
| C1262477 | Weight decreased | -0.0648 | 22.79% | 36.52% | 10.08% |
| C0168634 | BaseLine dental cement | -0.0677 | 35.58% | 52.17% | 20.23% |
| RXNORM:17767 | Amlodipine | -0.0761 | 16.79% | 24.65% | 9.52% |
| C0009676 | Confusion | -0.0817 | 43.53% | 60.50% | 27.82% |
| C0004093 | Asthenia | -0.0906 | 32.95% | 51.96% | 15.35% |
| C0042845 | Vitamin B 12 | -0.1030 | 28.20% | 42.06% | 15.37% |
| C5425799 | All other | -0.1042 | 47.99% | 67.57% | 29.85% |
| C4534363 | At home | -0.1405 | 46.55% | 69.16% | 25.62% |
| C0595998 | Household composition | -0.1820 | 49.69% | 73.23% | 27.89% |

Note: Feature importance was calculated by the difference in mean adjusted feature intensity between the clusters. Features with positive feature importance values were associated with Group 1, whereas features with negative feature importance values are associated with Group 2.

**S-Table 6.** Importance and prevalence of AD-related features included in the clustering model one year after AD diagnosis.

| Feature | Description | Feature Importance | Prevalence | | |
|---|---|---|---|---|---|
| | | | Overall | Group 1 | Group 2 |
| RXNORM:6719 | Memantine | 0.2194 | 34.99% | 39.05% | 31.24% |
| RXNORM:135447 | Donepezil | 0.1926 | 59.24% | 58.59% | 59.84% |
| PheCode:290.1 | Dementias | 0.1591 | 98.65% | 98.14% | 99.11% |
| PheCode:290 | Delirium dementia and amnestic and other cognitive disorders | 0.1570 | 98.66% | 98.16% | 99.11% |
| PheCode:332 | Parkinson's disease | 0.0862 | 5.57% | 8.60% | 2.78% |
| RXNORM:183379 | Rivastigmine | 0.0801 | 11.24% | 13.82% | 8.86% |
| PheCode:290.11 | Alzheimer's disease | 0.0753 | 98.37% | 97.62% | 99.06% |
| C0030567 | Parkinson disease | 0.0745 | 7.60% | 9.99% | 5.40% |
| RXNORM:103990 | Carbidopa/levodopa | 0.0575 | 3.53% | 5.99% | 1.25% |
| C0242422 | Parkinsonian disorders | 0.0187 | 4.07% | 5.54% | 2.71% |
| RXNORM:77492 | Tamsulosin | 0.0145 | 12.66% | 9.82% | 15.28% |
| C0025242 | Memantine | 0.0121 | 17.76% | 15.73% | 19.64% |
| C0011265 | Presenile dementia | 0.0048 | 84.64% | 77.68% | 91.07% |
| RXNORM:61381 | Olanzapine | 0.0026 | 5.27% | 3.89% | 6.55% |
| RXNORM:25025 | Finasteride | 0.0022 | 4.93% | 4.28% | 5.52% |
| PheCode:292 | Neurological disorders | 0.0019 | 61.28% | 50.70% | 71.07% |
| RXNORM:4637 | Galantamine | 0.0010 | 1.37% | 1.48% | 1.27% |
| PheCode:295 | Schizophrenia and other psychotic disorders | 0.0005 | 11.83% | 9.24% | 14.22% |
| RXNORM:1791685 | Pimavanserin | 0.0001 | 0.09% | 0.17% | 0.02% |
| C0149843 | Punch drunk syndrome | 0.0000 | 0.01% | 0.02% | 0.00% |
| C0751587 | CADASIL Syndrome | 0.0000 | 0.12% | 0.11% | 0.12% |
| C0016957 | Galactosylceramidase | 0.0000 | 0.00% | 0.00% | 0.00% |
| C0393483 | Brainstem encephalitis | 0.0000 | 0.00% | 0.00% | 0.00% |
| C4763868 | Cocaine use disorder | 0.0000 | 0.02% | 0.00% | 0.03% |
| C0238111 | Lennox-Gastaut syndrome | 0.0000 | 0.01% | 0.00% | 0.02% |
| C3662068 | Static encephalopathy | 0.0000 | 0.03% | 0.00% | 0.05% |
| C0260942 | Encounter due to screening for depression | 0.0000 | 0.22% | 0.17% | 0.26% |
| C0085220 | Cerebral amyloid angiopathy | 0.0000 | 0.30% | 0.19% | 0.40% |
| PheCode:333.3 | Tics and choreas | -0.0001 | 0.07% | 0.02% | 0.12% |
| C0041296 | Tuberculosis | -0.0003 | 1.28% | 0.73% | 1.79% |
| C3714756 | Intellectual disability | -0.0014 | 1.24% | 0.71% | 1.74% |
| C4484264 | Clark levels | -0.0018 | 2.98% | 1.95% | 3.92% |
| C0011127 | Pressure ulcer | -0.0018 | 3.58% | 1.91% | 5.12% |
| C0123091 | Quetiapine | -0.0019 | 4.48% | 2.55% | 6.27% |

| | | | | | |
|---|---|---|---|---|---|
| PheCode:430.2 | Intracerebral hemorrhage | -0.0020 | 1.52% | 0.62% | 2.34% |
| PheCode:818 | Intracranial hemorrhage (injury) | -0.0037 | 2.32% | 1.11% | 3.44% |
| C0007820 | Cerebrovascular disorders | -0.0037 | 5.64% | 2.95% | 8.13% |
| C0011269 | Dementia, vascular | -0.0048 | 6.31% | 3.77% | 8.66% |
| C0022983 | Laminectomy | -0.0052 | 4.05% | 2.18% | 5.78% |
| C0003537 | Aphasia | -0.0071 | 6.36% | 3.36% | 9.13% |
| PheCode:430 | Intracranial hemorrhage | -0.0073 | 3.51% | 1.75% | 5.14% |
| RXNORM:43611 | Latanoprost | -0.0079 | 4.43% | 2.87% | 5.87% |
| RXNORM:35636 | Risperidone | -0.0097 | 7.25% | 5.95% | 8.46% |
| PheCode:433.8 | Late effects of cerebrovascular disease | -0.0099 | 5.50% | 2.03% | 8.72% |
| C0002395 | Alzheimer's disease | -0.0100 | 83.32% | 76.65% | 89.49% |
| PheCode:345 | Epilepsy, recurrent seizures, convulsions | -0.0101 | 9.00% | 6.06% | 11.72% |
| PheCode:350.2 | Abnormality of gait | -0.0104 | 21.99% | 15.37% | 28.11% |
| PheCode:292.4 | Altered mental status | -0.0105 | 28.96% | 20.05% | 37.19% |
| C0008845 | Citalopram | -0.0109 | 7.43% | 4.26% | 10.37% |
| RXNORM:114477 | Levetiracetam | -0.0111 | 4.90% | 2.67% | 6.96% |
| RXNORM:221147 | Polyethylene glycol 3350 | -0.0118 | 18.46% | 11.08% | 25.30% |
| C0518460 | Bathing self care | -0.0126 | 10.92% | 5.80% | 15.66% |
| C0011168 | Deglutition disorders | -0.0132 | 13.05% | 6.91% | 18.74% |
| RXNORM:321988 | Escitalopram | -0.0141 | 9.68% | 7.38% | 11.81% |
| PheCode:591 | Urinary tract infection | -0.0145 | 29.33% | 21.10% | 36.95% |
| C0278061 | Abnormal mental state | -0.0153 | 21.01% | 12.80% | 28.60% |
| C0019360 | Herpes zoster disease | -0.0158 | 12.10% | 7.02% | 16.79% |
| RXNORM:51272 | Quetiapine | -0.0165 | 11.33% | 8.71% | 13.75% |
| C0007787 | Transient ischemic attack | -0.0202 | 13.02% | 7.32% | 18.28% |
| C0430533 | Dental diagnostic procedure | -0.0237 | 19.95% | 10.76% | 28.46% |
| C0428977 | Bradycardia | -0.0242 | 14.23% | 7.79% | 20.18% |
| RXNORM:15996 | Mirtazapine | -0.0245 | 9.25% | 6.25% | 12.03% |
| RXNORM:10737 | Trazodone | -0.0252 | 9.45% | 6.01% | 12.64% |
| PheCode:433.21 | Cerebral artery occlusion, with cerebral infarction | -0.0282 | 11.00% | 5.26% | 16.32% |
| C0527316 | Donepezil | -0.0377 | 30.91% | 22.86% | 38.36% |
| C0011053 | Deafness | -0.0404 | 20.57% | 12.77% | 27.78% |
| PheCode:798 | Malaise and fatigue | -0.0406 | 37.10% | 24.57% | 48.69% |
| C0034991 | Rehabilitation therapy | -0.0458 | 23.18% | 11.02% | 34.43% |
| RXNORM:36437 | Sertraline | -0.0473 | 15.71% | 11.51% | 19.60% |
| C0042029 | Urinary tract infection | -0.0528 | 25.16% | 14.42% | 35.09% |
| C0757844 | TNFSF13 protein, human | -0.0624 | 31.67% | 17.42% | 44.85% |

| | | | | | |
|---|---|---|---|---|---|
| C0206275 | Widowhood | -0.0625 | 16.80% | 8.49% | 24.48% |
| PheCode:433.3 | Cerebral ischemia | -0.0633 | 35.31% | 23.47% | 46.28% |
| C3809991 | Immunodeficiency common variable, 10 | -0.0680 | 26.37% | 14.29% | 37.54% |
| C1856053 | Hydranencephaly with renal aplasia-dysplasia | -0.0710 | 32.28% | 17.33% | 46.12% |
| RXNORM:11248 | Cyanocobalamin | -0.0738 | 22.19% | 14.21% | 29.57% |
| C1262477 | Weight decreased | -0.0764 | 31.96% | 19.95% | 43.06% |
| C4042877 | Clinical decision-making | -0.0874 | 52.18% | 34.48% | 68.55% |
| C0042571 | Vertigo | -0.0886 | 36.95% | 22.51% | 50.30% |
| PheCode:433 | Cerebrovascular disease | -0.0983 | 44.20% | 32.33% | 55.18% |
| C0439044 | Living alone | -0.1022 | 28.59% | 14.77% | 41.36% |
| C0034770 | Mental recall | -0.1065 | 51.81% | 34.41% | 67.91% |
| RXNORM:17767 | Amlodipine | -0.1265 | 23.35% | 15.37% | 30.73% |
| C0009676 | Confusion | -0.1273 | 58.89% | 43.27% | 73.35% |
| C0168634 | BaseLine dental cement | -0.1315 | 50.72% | 31.88% | 68.15% |
| C0038454 | Cerebrovascular accident | -0.1366 | 35.72% | 20.71% | 49.61% |
| C0004093 | Asthenia | -0.1435 | 44.88% | 24.87% | 63.38% |
| C5425799 | All other | -0.1476 | 61.91% | 42.80% | 79.58% |
| C0042845 | Vitamin B 12 | -0.1610 | 38.65% | 25.89% | 50.46% |

Note: Feature importance was calculated by the difference in mean adjusted feature intensity between the clusters. Features with positive feature importance values were associated with Group 1, whereas features with negative feature importance values are associated with Group 2.

**S-Table 7**. Demographic and clinical profile of the overall study population with and without imputation of missing race and ethnicity.

| | With imputation of race and ethnicity [a] | Without imputation of race and ethnicity | P-value |
|---|---|---|---|
| **Number of Patients, N (%)** | 16411 (100%) | 15,618 (100%) | |
| **Age at AD diagnosis, mean (SD), years** | 81.18 (8.98) | 81.19 (8.95) | >0.9 |
| **Gender, N (%)** | | | >0.9 |
| Women | 10,559 (64%) | 10,053 (64%) | |
| Men | 5852 (36%) | 5,565 (36%) | |
| **Race/ethnicity, N (%)** | | | >0.9 |
| Non-Hispanic White | 15,105 (92%) | 14,381 (92%) | |
| American Indian | 8 (<0.1%) | 7 (<0.1%) | |
| Asian | 65 (0.4%) | 59 (0.4%) | |
| Black | 1,183 (7.2%) | 1,129 (7.2%) | |
| Hawaiian or Pacific Islander | 3 (<0.1%) | 2 (<0.1%) | |
| Hispanic or Latino | 47 (0.3%) | 40 (0.3%) | |
| **Elixhauser Comorbidity Index [b], mean (SD)** | 9.04 (9.52) | 9.08 (9.54) | >0.9 |
| **Baseline comorbidities (top 2 prevalent), N (%)** | | | |
| Hypertension, uncomplicated | 12,052 (73%) | 11,507 (74%) | 0.6 |
| Cardiac arrhythmia | 6,326 (39%) | 6,051 (39%) | 0.7 |
| **AD-related medication [c], N (%)** | | | |
| Not prescribed | 3,709 (23%) | 3,478 (22%) | 0.5 |
| Prescribed | 12,702 (77%) | 12,140 (78%) | |
| Before AD diagnosis | 8,885 (70%) | 8,493 (70%) | >0.9 |
| On or after AD diagnosis | 3,817 (30%) | 3,647 (30%) | |
| **Follow-up duration, median months (1st and 3rd quantiles)** | 78 (48, 109) | 78 (49, 110) | >0.9 |

Note:

a. We imputed missing race and ethnicity based on age and gender.
b. We calculated the comorbidity burden using 24 months of pre-index data (*i.e.,* in the two years preceding AD diagnosis).
c. Please see S-Table 4 for the list of AD-related medications.

**S-Table 8.** Baseline demographic and clinical profile of AD patient groups clustered one year after AD diagnosis.

| | Overall | Group 1 | Group 2 | P-value |
|---|---|---|---|---|
| **Number of Patients, N (%)** | 12606 (100%) | 5759 (45.7%) | 6847 (54.3%) | |
| **Age at AD diagnosis, mean (SD)** | 80.4 (9.1) | 80.8 (9.4) | 80.1 (8.8) | **<.0001** |
| **Gender, N (%)** | | | | .473 |
| Women | 8161 (64.7%) | 3748 (65.1%) | 4413 (64.5%) | |
| Men | 4445 (35.3%) | 2011 (34.9%) | 2434 (35.5%) | |
| **Race/ethnicity [a], N (%)** | | | | **<.001** |
| Non-Hispanic white | 11607 (92.1%) | 5240 (91.0%) | 6367 (93.0%) | |
| American Indian | 8 (0.1%) | 5 (0.1%) | 3 (0.0%) | |
| Asian | 59 (0.5%) | 35 (0.6%) | 24 (0.4%) | |
| Black | 893 (7.1%) | 460 (8.0%) | 433 (6.3%) | |
| Hawaiian or Pacific Islander | 2 (0.0%) | 0 (0.0%) | 2 (0.0%) | |
| Hispanic or Latino | 37 (0.3%) | 19 (0.3%) | 37 (0.3%) | |
| **Elixhauser Comorbidity Index [b], mean (SD)** | 8.2 (9.2) | 10.4 (10.0) | 6.4 (8.0) | **<.0001** |
| **Baseline comorbidities (top 2 prevalent), N (%)** | | | | **<.0001** |
| Hypertension, uncomplicated | 9061 (71.9%) | 4499 (78.1%) | 4562 (66.6%) | |
| Cardiac arrhythmias | 4483 (35.6%) | 2538 (44.1%) | 1945 (28.4%) | |
| **AD-related medication [c], N (%)** | | | | |
| Not prescribed | 2413 (19.1%) | 1310 (22.7%) | 1103 (16.1%) | **<.0001** |
| Prescribed | 10193 (80.9%) | 4449 (77.3%) | 5744 (83.9%) | |
| Before AD diagnosis | 5703 (45.3%) | 2759 (47.9%) | 2944 (43.0%) | **<.0001** |
| On or after AD diagnosis | 4490 (35.6%) | 1690 (29.3%) | 2800 (40.9%) | |
| **Follow-up months from baseline, median months (1st and 3rd quantiles)** | 80 (50, 111) | 63 (35, 86) | 88 (61, 126) | **<.0001** |

a. We imputed missing race and ethnicity based on age and gender.
b. We calculated the comorbidity burden using 24 months of pre-index data (*i.e.,* during the two years prior to AD diagnosis).
c. List of AD-related medications can be found in S-Table 4.

**S-Table 9.** Comparison of 2-, 5- and 10-year rate of nursing home admission and death for AD patient groups clustered one year after AD diagnosis (N=12606).

| **Time of Clustering** | | **Nursing Home Admission** | | | **Death** | | |
|---|---|---|---|---|---|---|---|
| | | **Group 1** | **Group 2** | **P-value** | **Group 1** | **Group 2** | **P-value** |
| **At AD diagnosis** | **2 years** | 14.9% (13.9%, 15.9%) | 11.6% (10.9%, 12.3%) | **<0.0001** | 10.4% (9.6%, 11.3%) | 8.9% (8.3%, 9.6%) | **0.00465** |
| | **5 years** | 34.6% (33.3%, 35.9%) | 32.5% (31.4%, 33.5%) | **0.0144** | 33.7% (32.4%, 35.0%) | 35.7% (34.7%, 36.8%) | **0.0192** |
| | **10 years** | 39.1% (37.8%, 40.5%) | 41.4% (40.3%, 42.5%) | **0.0107** | 40.3% (39.0%, 41.7%) | 52.7% (51.6%, 53.8%) | **<0.0001** |
| **1 year after AD diagnosis** | **2 years** | 14.6% (13.7%, 15.5%) | 11.5% (10.7%, 12.2%) | **<0.0001** | 11.0% (10.3%, 11.9%) | 8.2% (7.6%, 8.9%) | **<0.0001** |
| | **5 years** | 34.5% (33.3%, 35.8%) | 32.3% (31.2%, 33.4%) | **0.0082** | 35.1% (33.9%, 36.3%) | 34.8% (33.6%, 35.9%) | **<0.0001** |
| | **10 years** | 39.6% (38.3%, 40.9%) | 41.3% (40.1%, 42.5%) | 0.060 | 52.4% (51.2%, 53.6%) | 42.5% (41.2%, 43.7%) | **<0.0001** |

**S-Table 10.** Comparison of the demographic characteristics of the UPMC AD cohort with other EHR-based AD cohorts and a nationwide multisite prospective AD cohort.

| | UPMC | OneFlorida | UCSF | Mt Sinai | WCM | NACC |
|---|---|---|---|---|---|---|
| **Number of Patients, N** | 16411 | 29922 | 8804 | 5958 | 792 | 14044 |
| **Age at AD diagnosis, mean (SD), years** | 81.2 (9.0) | 79.87 (9.66) [a] | 86.5 (6.4) [b] | 88.3 (8.7) [b] | 78.4 (5.4) | NR |
| **Age at AD symptom onset, median (IQR), years** | NR | NR | NR | NR | NR | 72 (64, 78) |
| **Women, N (%)** | 10559 (64.3%) | 20951 (70.0%) | 5558 (63.1%) | 4138 (69.5%) | 487 (61.5%) | 7850 (55.9%) |
| **Race, N (%)** | | | | | | |
| White | 15105 (92%) | 13065 (43.7%) | 5462 (64.0%) | 2904 (48.7%) | 484 (54.8%) | 11538 (82.2%) |
| American Indian | 8 (0.0%) | 36 (0.1%) | 9 (0.1%) | 8 (0.1%) | NR | NR |
| Asian | 65 (0.4%) | 368 (1.2%) | 879 (10.3%) | 78 (1.3%) | 15 (1.9%) | NR |
| Black | 1183 (7.2%) | 4599 (15.4%) | 586 (6.9%) | 1214 (20.4%) | 70 (8.8%) | 1653 (11.8%) |
| Hawaiian or Pacific Islander | 3 (0.0%) | 7 (0%) | 452 (5.3%) | 5 (0.1%) | NR | NR |
| Other / Unknown | 0 (0%) | 11847 (39.6%) | 1416 (16.1%) | 1749 (29.4%) | 273 (34.5%) | 853 (6.1%) |
| **Hispanic or Latino, N (%)** | 47 (0.3%) | NR | NR | NR | NR | 1241 (8.8%) |

Abbreviations: UCSF - University of California San Francisco, WCM - Weill Cornell Medicine, NACC - National Alzheimer's Coordinating Center, NR - not reported.

Note:

a. Since only the patient counts for different age ranges at AD diagnosis (*e.g.,* <65, 65-74, 74-85, ≥85) were provided, the mean and standard deviation were estimated using grouped data. We assigned midpoints to each age range (*e.g.,* the midpoint of 65-74 age range was 69.5; the midpoint of 75-84 age range was 79.5). For open-ended intervals, representative midpoints were assumed (*e.g.,* the midpoint for <65 was 59.5; the midpoint for ≥85 was 89.5). We then applied frequency-weighted formulas to calculate the approximate mean and standard deviation.

b. Estimated age is included since dates are shifted (by less than a year) during the de-identification process.

**S-Table 11.** Number of patients with high healthcare utilization (n=7713) in each baseline cluster.

| | | Clustering of Patients with High Healthcare Utilization (Stratified Analysis) | | |
|---|---|---|---|---|
| | | Group 1 | Group 2 | Total |
| Clustering of All Patients (Main Analysis) | Group 1 | 4065 | 652 | 4717 |
| | Group 2 | 727 | 2269 | 2996 |
| | Total | 4792 | 2921 | 7713 |

**S-Table 12.** Number of patients with low healthcare utilization group (n=8698) in each baseline cluster.

| | | Clustering of Patients with Low Healthcare Utilization (Stratified Analysis) | | |
|---|---|---|---|---|
| | | Group 1 | Group 2 | Total |
| Clustering of All Patients (Main Analysis) | Group 1 | 1514 | 503 | 2017 |
| | Group 2 | 821 | 5860 | 6681 |
| | Total | 2335 | 6363 | 8698 |

### SUPPLEMENTARY FIGURES

**S-Figure 1.** (A) Graphical representation of Poisson LogNormal Mixture Model (PLMM). Squares indicate model parameters; circles indicate random variables; Filled-in shapes indicate observed variables. The indication [p] means a vector of size p. (B) Underlying model assumptions. We assumed a generative modeling framework in which healthcare utilization (U) affects the cluster membership Y (bottom layer). The healthcare utilization and cluster membership together generate the latent low-rank patient representation (Z) through guided projection of electronic health records (EHR) feature embeddings (middle layer). The observed EHR feature counts (X) are generated from the low-rank patient embeddings (top layer). The effect of cluster membership (Y) and healthcare utilization (U) on observed EHR feature counts (X) is fully captured by the latent layer of patient representations (Z).

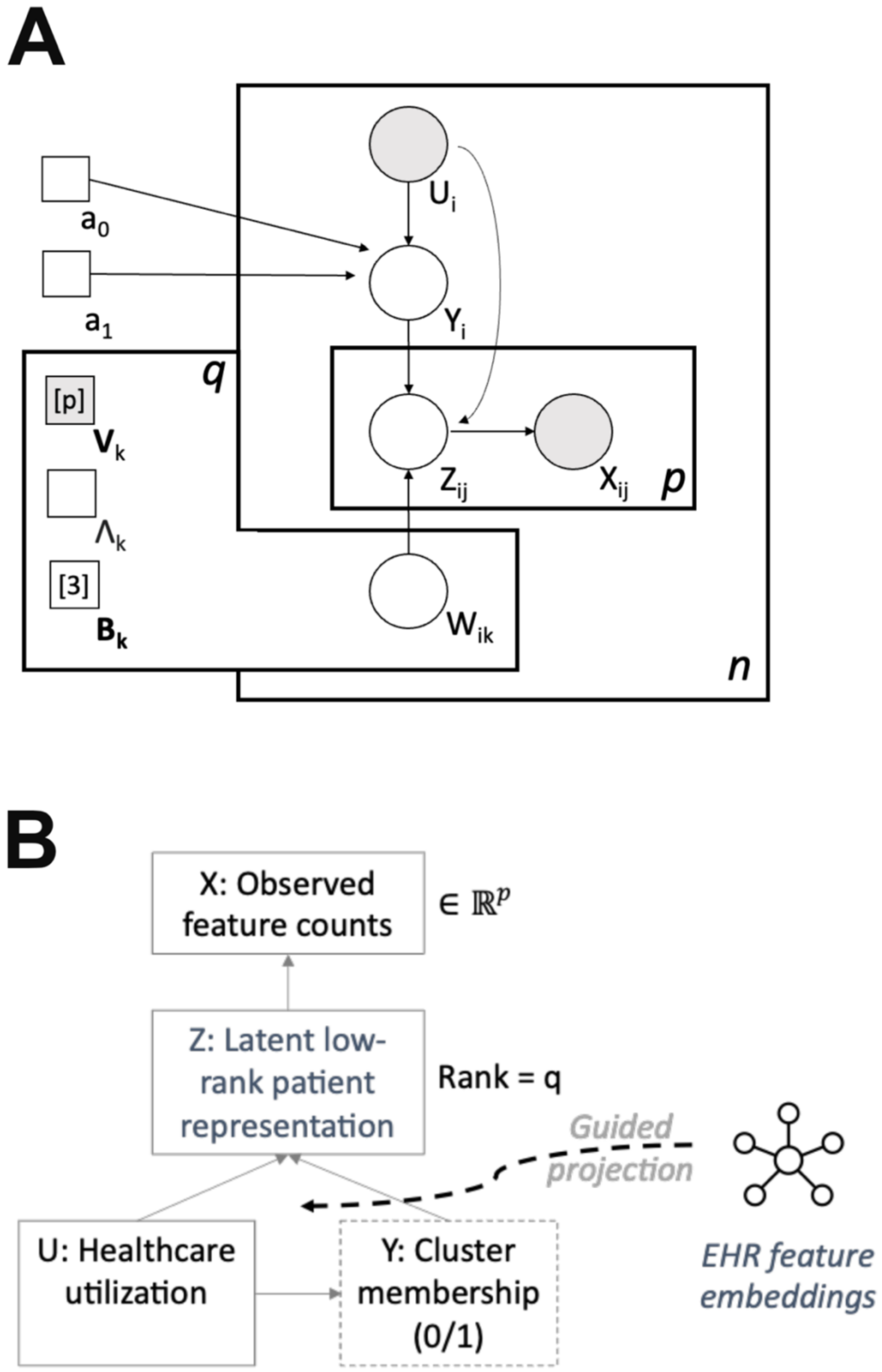
A
$a_0$
$a_1$
$U_i$
$Y_i$
$Z_{ij}$
$X_{ij}$
p
q
[p]
$\mathbf{V}_k$
$\Lambda_k$
[3]
$\mathbf{B}_k$
$W_{ik}$
n
B
X: Observed feature counts
$\in \mathbb{R}^p$
Z: Latent low-rank patient representation
Rank = q
Guided projection
U: Healthcare utilization
Y: Cluster membership (0/1)
EHR feature embeddings

**S-Figure 2.** Baseline comorbidity burden of AD patient groups clustered one year after AD diagnosis.

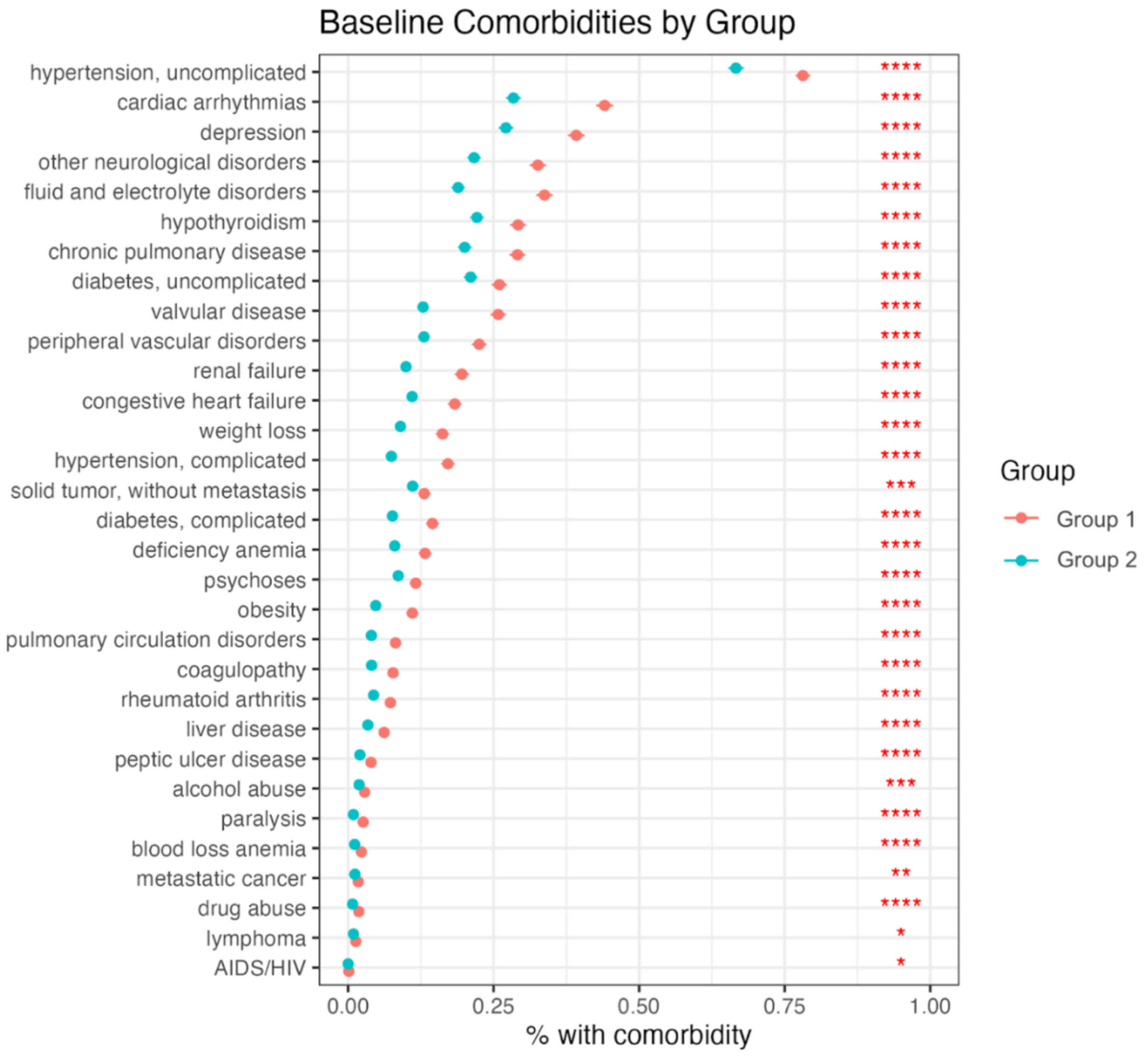

**S-Figure 3.** Adjusted survival curve of (A) time to nursing home and (B) time to death for AD patient groups clustered one year after AD diagnosis.

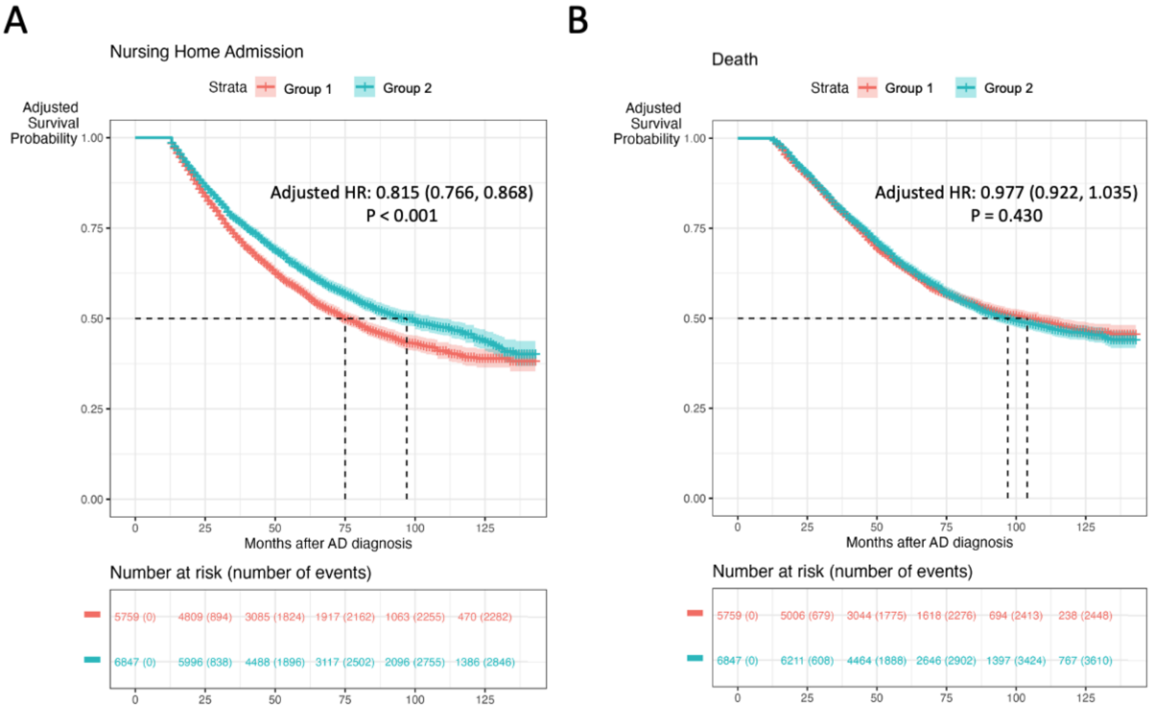

**S-Figure 4.** Top 15 features contributing to differences between AD patient groups clustered one year after AD diagnosis. Features in the color salmon were positively associated with Group 1 membership, while features in the color teal were negatively associated with Group 1 membership. (A) Word cloud of the important features. Text size represented the relative importance of the feature. (B) Correlation matrix of the features.

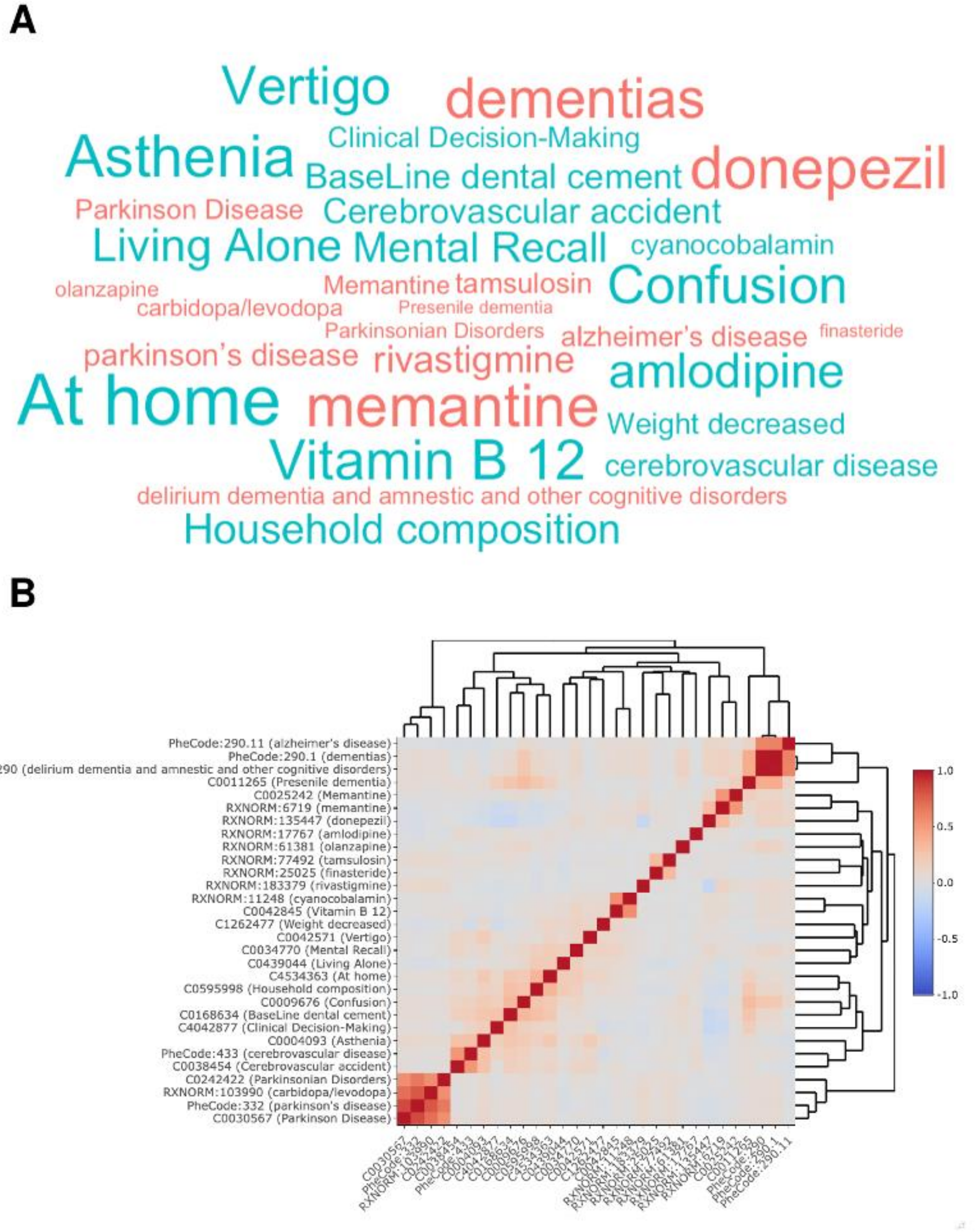

**S-Figure 5.** Adjusted survival curve of (A) time to nursing home and (B) time to death for AD patient groups clustered at AD diagnosis, with K=3.

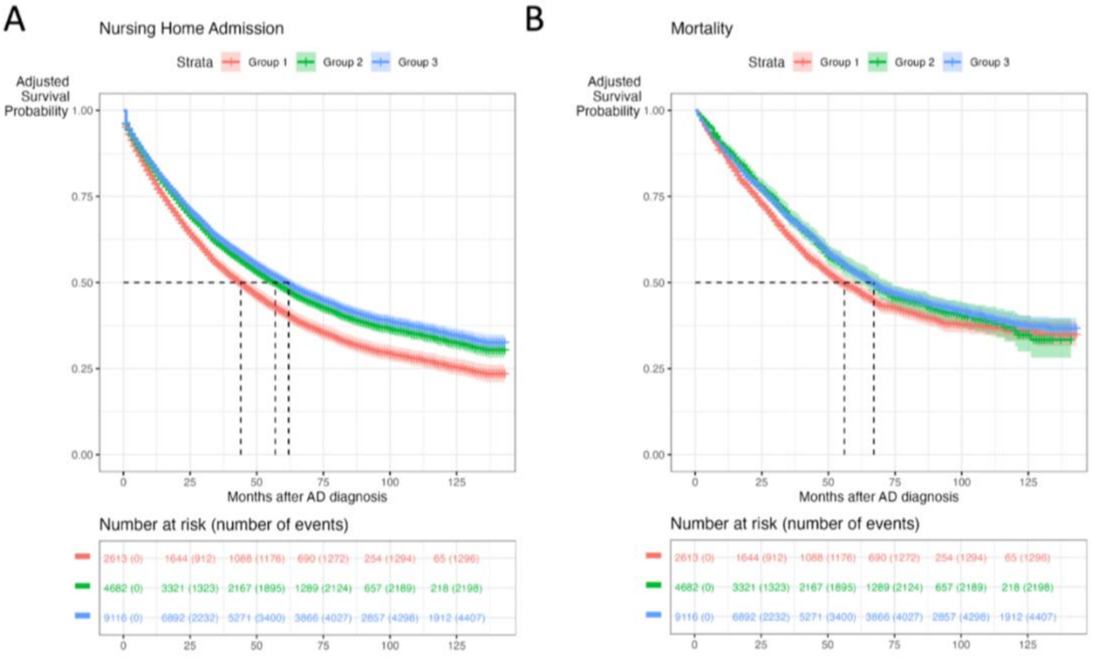

**S-Figure 6**. Adjusted survival curve of time to nursing home (A, C) and time to death (B, D) for AD patient groups clustered at AD diagnosis using benchmark methods K-means clustering (A-B) and Gaussian Mixture Model (C-D).

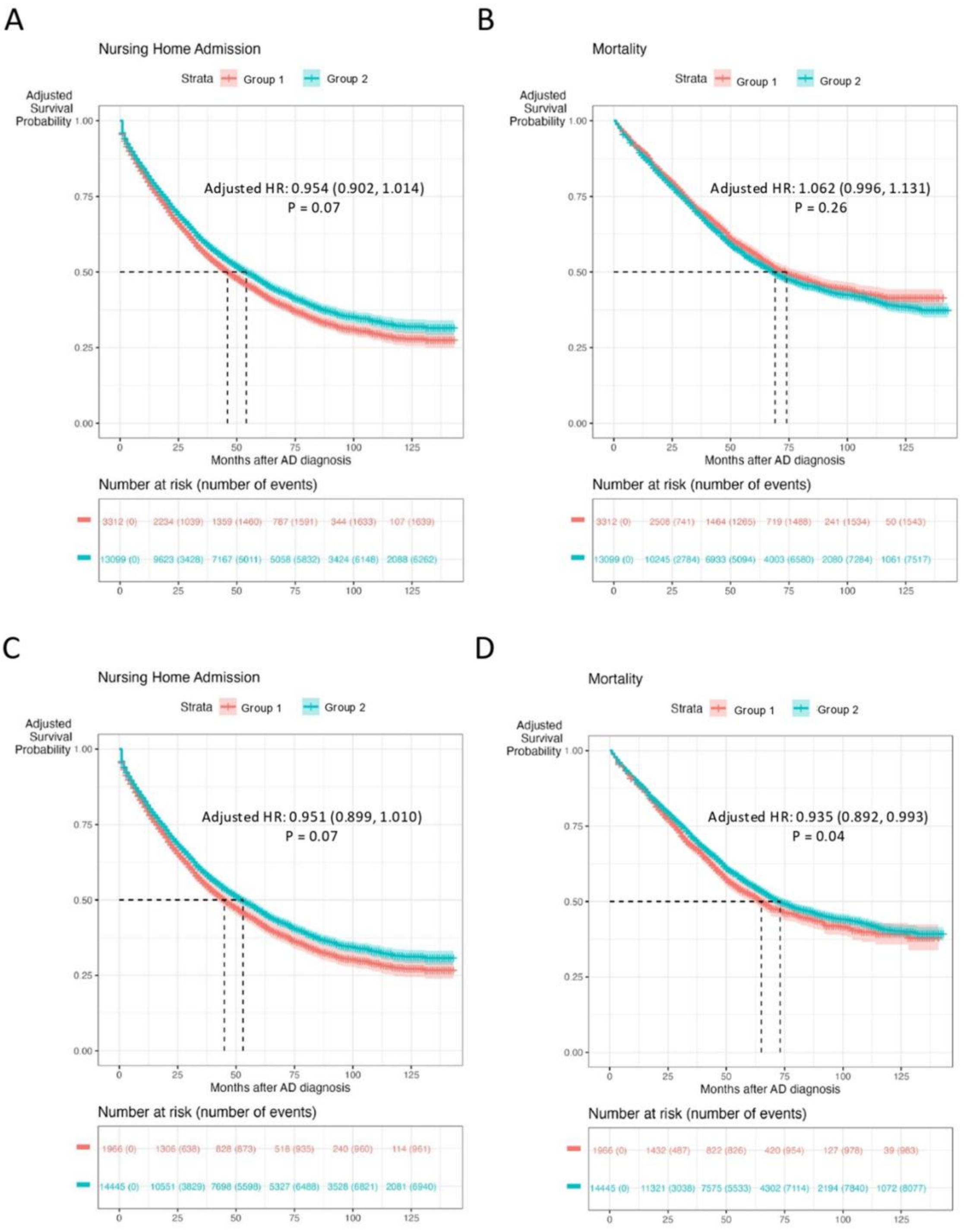
